# A tuneable structural switch controls RNF213 activity and is bypassed in Moyamoya disease

**DOI:** 10.64898/2026.10.07.26364583

**Authors:** Lillie E. Bell, Victoria Faas, Daniel B. Grabarczyk, Moritz H. Wallnöfer, Eliska Kohoutkova, Luiza Deszcz, Thomas L. Williams, Anton Meinhart, Harry Scott, Florence Riant, Chaker Aloui, Manoelle Kossorotoff, Laure Lacan, François Godart, Elisabeth Tournier-Lasserve, Adam J. Fletcher, Tim Clausen

## Abstract

RNF213 is a giant AAA+ ubiquitin ligase that serves as a central hub of innate immunity, responding to diverse stress signals with broad reactivity^1-6^. This versatility demands tight regulation, and its failure causes disease, such as Moyamoya angiopathy^7-10^ and cancer progression^11-13^. How the mega-enzyme is restrained and how mutations override this control is unknown. Here, we show that RNF213 activity is regulated by a dimeric switch — a constitutive dimer that alternates between auto-inhibited and activation-competent states. In the closed dimer, intermolecular contacts bury the E3 machinery, whereas ATP binding remodels the AAA+ core to open the dimer and prime ubiquitination. Dimer conversion is tuned by ubiquitination factors, redox conditions and post-translational modifications, allowing multiple stress signals to act through the same activation mechanism. Moyamoya mutations subvert the same open-closed transition, destabilizing the autoinhibited dimer in a graded manner that tracks with clinical severity. Structural, biochemical and cellular data reveal how disease mutations allosterically disrupt the dimer interface and bypass ATP-dependent control to drive NF-κB activation and caspase-dependent apoptosis. Together, our findings reveal a tuneable dimeric switch that converts diverse stressors into graded inflammatory signalling and provide a mechanistic link between genetic predisposition, environmental triggers and Moyamoya disease.

---

Ubiquitin-dependent signalling is a central mechanism by which eukaryotic cells sense stress and coordinate inflammatory and innate immune responses. A key node within this network is the NF-κB family of transcription factors, which integrates inputs from infection, metabolic stress and organelle damage to control cellular fate decisions^14,15^. Since inappropriate activation of NF-κB pathways can drive hyperinflammation, cell death and disease, the ubiquitin ligases that regulate them must also be subject to tight control.

RNF213 has emerged as a key regulator of NF-κB signalling. The giant AAA+ ubiquitin ligase responds to diverse cellular stresses^4,16^ and promotes inflammatory signalling through ubiquitination of the CYLD–SPATA2 complex, thereby relieving a brake on LUBAC-dependent NF-κB activation^11,17,18^. RNF213 also plays direct roles in host defence by targeting intracellular pathogens. It catalyses ubiquitination of bacterial lipids, marking pathogen-associated membranes for downstream immune responses and autophagic clearance^2,3,5,6,19^. Consistent with its central role in inflammatory signalling, RNF213 variants have been linked to diverse disease conditions, including vascular disorders such as coronary artery disease and pulmonary arterial hypertension^20,21^, as well as cancer, where RNF213 is recurrently mutated and amplified across tumour types^11,12^.

On a molecular level, RNF213 is unique among E3 ligases for targeting diverse protein and non-protein substrates and for being regulated by ATP levels in the cell that rise during infection and cellular stress^1^. The shift from resting to active RNF213 is thought to involve changes in oligomeric state. RNF213 forms higher molecular weight species upon interferon stimulation^6^, coats pathogens through cooperative recruitment^22^ and undergoes activity-dependent assembly changes during inflammatory signalling^11^. Together, these observations suggest that changes in oligomeric states represent a fundamental layer of RNF213 regulation. However, despite extensive structural and biochemical studies^1,22-25^, it remains unclear how changes in RNF213 assembly affect the activity of the E3 ligase.

Understanding how RNF213 is regulated is especially important for Moyamoya angiopathy (MMA), a severe vascular disorder strongly linked to single-nucleotide polymorphisms in RNF213^9,10^, many of which cluster within its E3 module^8^. How these disease mutations perturb RNF213 activity is unknown. Here, we show that RNF213 acts not as a monomeric ligase, as commonly assumed, but as a constitutive dimer that switches between distinct self-assembly states to control its activity. We define open and closed dimeric assemblies that correspond to active and inactive E3 states, uncover how ATP binding, redox chemistry and ubiquitination factors drive dimer interconversion and show how disease-associated mutations can destabilise the inactive state. These findings establish dimerization-dependent control as the core principle of RNF213 regulation and explain how Moyamoya mutations bypass an intrinsic safeguard mechanism to dysregulate inflammatory signalling.

## RESULTS

### RNF213 forms a constitutive dimer that transitions between open and closed states

To define the molecular basis of RNF213 oligomer formation and its impact on activity, we first expressed the human orthologue and separated distinct assembly states by size-exclusion chromatography (SEC) (**Figure 1A; left**). Mass photometry^26^ revealed two major populations: a low-molecular-weight (LMW) species with an apparent mass of ∼600 kDa and a high-molecular-weight (HMW) species of ∼1200 kDa (**Figure 1A; right**), consistent with complexes containing either one or two RNF213 cores.

**Figure 1:**
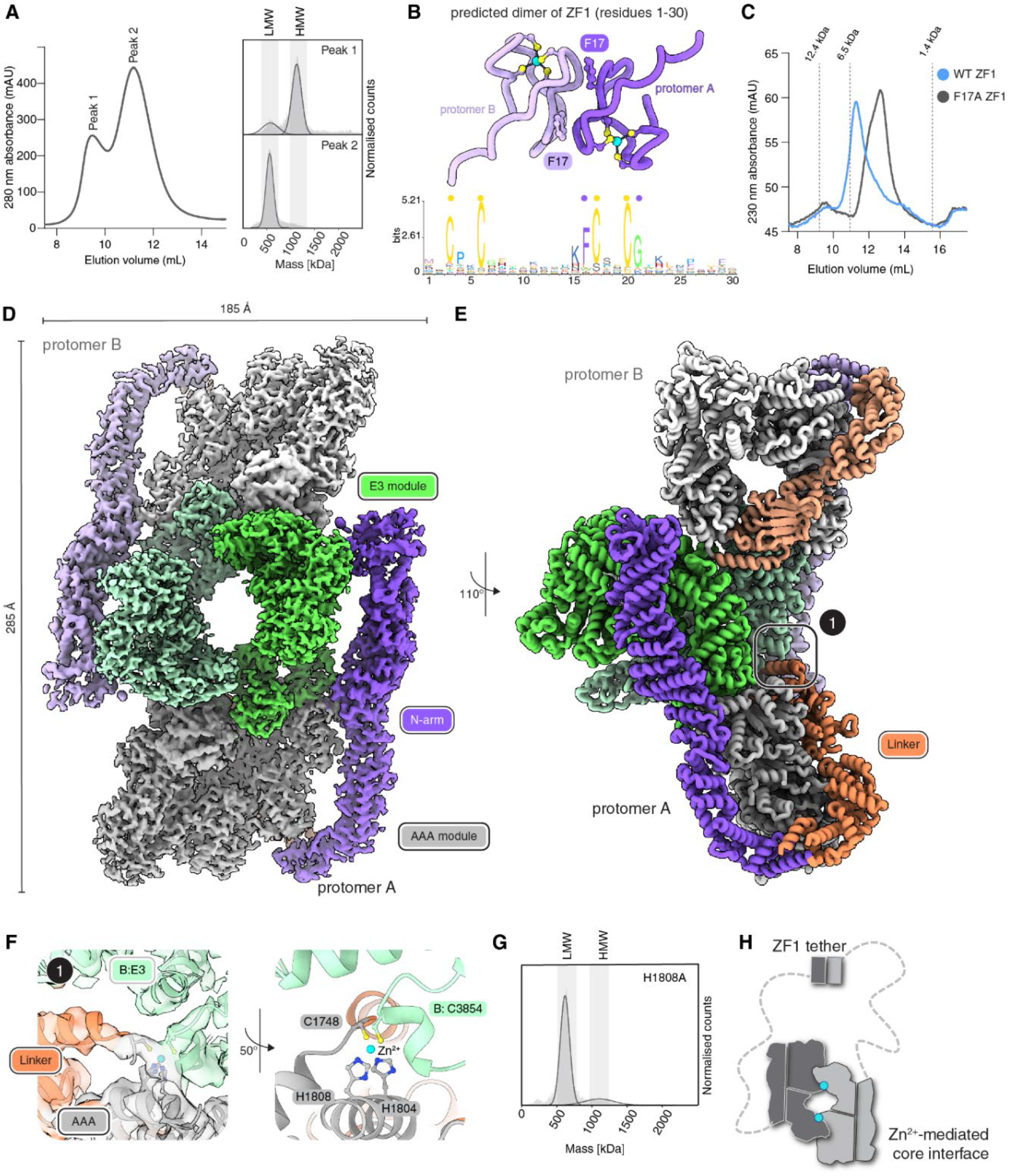
RNF213 forms a constitutive dimer that transitions between open and closed states. **(A)** SEC analysis of WT RNF213 with two distinct peaks labelled 1 and 2 respectively. The right panel shows the mass photometry analysis of SEC peaks 1 and 2. **(B)** AlphaFold3 prediction of the ZF1 dimer, with the two N-terminal segments (residues 1-30) coloured in different shades of purple. The plot below shows the vertebrate conservation of residues 1-30. Zinc-coordinating (yellow dots) and dimer interface residues (purple dots) are indicated. **(C)** SEC analysis of WT (blue) and F17A (grey) ZF1 domain, with indicated size markers from SEC standards. **(D)** Composite cryo-EM map of the human RNF213 dimer coloured by module, with protomer B shown in lighter shades. **(E)** Side view of the RNF213 dimer model with box-1 highlighting the center of the core interface. **(F)** Zoom-in of box-1 showing the EM density of involved residues and the organization of the shared Zn^2+^ binding site. Contributed domains and residues by monomer B are labelled. **(G)** Mass photometry of H1808A mutant. **(H)** Schematic presentation of the RNF213 dimer, showing the two distinct interfaces formed by ZF1 and the protein core.

Mass spectrometry confirmed that both fractions contained RNF213 as the sole protein component. However, peptides derived from the extreme N-terminus (residues 1–30) were specifically depleted in the LMW fraction (**Figure S1A**), suggesting that this region is buried and thus protected upon HMW formation. Prediction of RNF213 intermolecular contacts using AlphaFold3^27^ identified a single high-confidence interaction within the first zinc finger (ZF1; residues 1–30) at the extreme N-terminus of the protein (**Figure 1B, Figure S1B**). The ZF1-ZF1 interface is centred on a hydrophobic surface surrounding the highly conserved residue Phe17. ZF1 alone formed a stable dimer in solution, as validated by SEC and native mass spectrometry (**Figure 1C, Figure S1C**). Even at nanomolar concentrations, monomeric species were not detected by SEC, indicating a high-affinity interaction. Substitution of the conserved Phe17 (F17A) abolished dimer formation of the isolated domain. In the context of full-length RNF213, however, deletion of ZF1 reduced but did not eliminate formation of the HMW species (**Figure S1D**), indicating the existence of an additional dimerization interface beyond ZF1.

To define the architecture of the HMW assembly, we determined its structure by cryo-electron microscopy (cryo-EM). Reconstruction at 3.1 Å resolution revealed a symmetric dimer in which two RNF213 protomers are aligned face-to-face (**Figure 1D, Figure S2A**). Density was well resolved for all structured regions (residues 375-5207), including the N-arm, linker region, dynein-like AAA module and E3 ligase region; only the catalytic RZ domain was poorly defined (**Figure S2B**). The two protein cores interact along their longitudinal axis through contacts between the AAA module of one protomer and the E3 module of the other (**Figure 1E**). At the centre of this core-core interface, an extended loop (residues 3845–3860) of the E3-back domain engages the AAA1 domain (residues 1735–1755) of the neighbouring protomer (**Figure 1F**) and hence is well positioned to sense nucleotide-dependent rearrangements within the adjacent AAA core.

Unexpectedly, the cryo-EM map also revealed two Zn^2+^ ions positioned within this E3-back/AAA1 interface (**Figure S2C**). Each Zn^2+^ ion is coordinated by three residues from one protomer (Cys1748, His1804 and His1808) and one residue from the opposing protomer (Cys3854), yielding two shared Zn^2+^-binding sites that bridge the RNF213 cores (**Figure 1F**). Consistent with a role in self-association, mutation of the Zn^2+^ coordinating residue His1808 (H1808A) strongly destabilized the HMW assembly and shifted RNF213 almost entirely into the LMW state (**Figure 1G**).

Together, these findings reveal that RNF213 dimerization is mediated through two mechanistically distinct interfaces. A high-affinity ZF1–ZF1 interaction forms a constitutive tether that links two RNF213 protomers, whereas a Zn^2+^-stabilized interface between the AAA and E3 regions aligns the protein cores into a compact assembly. The flexible

∼350-residue linker separating ZF1 from the structured protein body allows these interfaces to engage independently, enabling RNF213 to adopt two distinct dimeric architectures: a compact “closed dimer” (HMW in mass photometry), in which both interfaces are engaged, and an extended “open dimer” (LMW), in which the protein cores disengage and appear as separate particles, while remaining tethered via the ZF1 motif (**Figure 1H**). Thus, the functional unit of RNF213 is not a monomer, but a dimer that transitions between alternative assembly states. The broad conservation of the ZF1 tether across vertebrates suggests that dimeric assembly is an ancestral feature of this giant E3.

### ATP binding opens the closed dimer to prime the E3 module

To safeguard against its extreme substrate promiscuity, which extends to lipids, glycans, and protein N-termini **(Figure S3A)**, RNF213 is strictly gated by ATP, a metabolite whose levels rise during IFN signalling – a hallmark of pathogen detection ^1^. Yet, prior structural analyses could not resolve how nucleotide binding to the AAA module exerts control over the distant E3 module^1,22,24,25,28,29^. The closed dimer identified here offers a mechanistic explanation: the core-core interface between opposing protomers buries the catalytic RZ domain and the E2-binding CTD, thereby occluding the ubiquitination machinery (**Figure 2A**). We therefore explored whether nucleotide binding releases this autoinhibited architecture by shifting RNF213 from a closed into an open dimer, where the core-core interface is released. To test this model, we monitored dimer distribution over time by mass photometry, in which the open and closed dimers can be distinguished by their apparent molecular masses of ∼600 and ∼1200 kDa, respectively (**Figure 1A**). Incubation with the slowly hydrolysable analogue ATPγS induced a shift from the closed to the open population within ∼15 minutes, followed by re-formation of the closed dimer at later times (**Figure 2B**). This reversible response indicated that nucleotide binding transiently shifts the open-closed equilibrium, increasing the fraction of activation-competent dimers on a physiologically relevant timescale.

**Figure 2:**
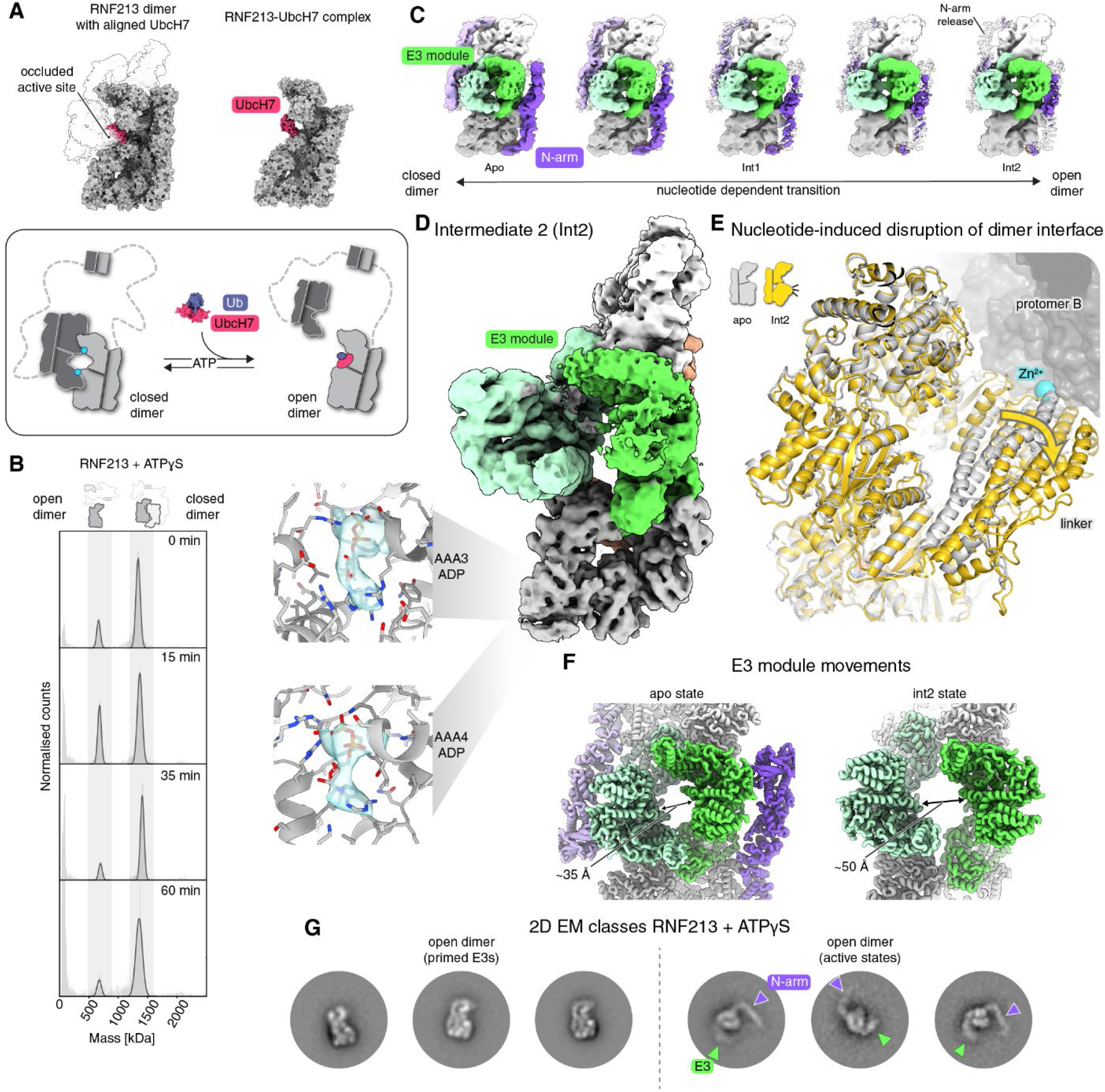
ATP binding opens the RNF213 dimer to expose its E3 active site. **(A)** Top panel: left, PDB: 7OIK (right) aligned with dimer protomer A to position UbcH7 on the dimeric complex. The alignment shows that protomer B (transparent) occludes the E2 binding position, sterically preventing interaction with UbcH7. Right, cryo-EM structure showing the binding mode of the E2 enzyme UbcH7 (magenta) to the RNF213 protomer (PDB: 7OIK). Bottom panel: Proposed model of how ATP-induced opening of the dimer would enable UbcH7∼Ub binding to promote E3 activity. **(B)** Time course mass photometry of RNF213 in the presence of ATPγS. **(C)** Snapshots across 3D variability trajectory reflecting structural changes of the open-closed transition (associated with **Movie S1**). Positions of Intermediate states 1 and 2 across the trajectory are indicated. **(D)** Cryo-EM density of second intermediate (Int2), where protomer B is twisted relative to A to promote E3 module separation. N-arm density is not observed in this state. ADP was bound in both AAA3 and AAA4 active sites (left panel). **(E)** Superposition of a single protomer of the closed dimer (grey) with a single protomer of Int2 (yellow), focusing on the AAA core and attached E3 and linker portions. The arrow indicates how reorientation of the AAA module leads to disruption of the core interface near the Zn^2+^ site. This is reflected by a >10 Å displacement of the helix-turn-helix segment harbouring two coordination sites. **(F)** E3 modules of apo vs nucleotide bound (int2) states displaying increased distance between the modules as a result of nucleotide binding. Measured distances between E3 modules of neighbouring protomers was carried out in ChimeraX. **(G)** Negative stain EM 2D classes of RNF213 + ATPγS incubation, showing the presence of primed states (left) and active states (right), the latter having characteristically blurred E3 modules and outward swung N-arm, as indicated by coloured arrows.

To visualize how nucleotide binding promotes dimer opening, we incubated RNF213 with ATPγS and collected cryo-EM data at early time points, marking the transition from resting to active states. Cryo-EM with 3D variability analysis captured a continuum of conformations bridging a compact closed dimer and gradually separated assemblies (**Figure 2C, Movie S1, Figure S4**). One intermediate class (int1; 3.4 Å) resembled the resting dimer but contained density for ADP in AAA3 (**Figure S3B)**, whereas a second prominent class (int2; 4.3 Å) contained nucleotides in the two functional AAA domains of RNF213, AAA3 and AAA4, and showed pronounced rearrangements in the AAA module (**Figure 2D**). Alignment of int2 with the resting state revealed an en bloc reorientation of the AAA1–AAA3 half relative to AAA4–AAA6, thereby distorting the core interface near AAA1 and inducing long-range conformational changes through the overarching linker domain (**Figure 2E; Figure S3C**). Resultant rearrangements propagated throughout the entire molecule, ultimately leading to the detachment of the N-arm and weakening of central interdimer contacts, as seen by an expansion of the gap between the two E3 modules from ∼35 Å to ∼50 Å at the center (**Figure 2F, Movie S2**). Less defined density for one protomer in these cryo-EM maps further confirmed separation of the two protein cores (**Movie S3**).

Although fully separated particles could not be resolved by cryo-EM, negative-stain EM of the open dimer sample readily distinguished two active RNF213 conformations, which were defined either by an aligned or a detached N-arm (**Figure 2G**). Detachment of the N-arm correlated with increased flexibility of the E3 module that appeared blurred in these particles. Since previously reported structures place the catalytic RZ domain and the CTD ∼25 Å apart, in a geometry unfavourable for ubiquitin transfer (**Figure S3D**), we infer that N-arm detachment and the resulting flexibility of the functional CTD and RZ motifs are likely necessary to enable transthiolation from E2∼Ub to RNF213, and hence that these particles represent the active state of the E3 ligase. Together, these results show how ATP binding drives large-scale conformational rearrangements that transmit local changes in the AAA core to the distant E3 module, opening the autoinhibited dimer into a ubiquitination-competent state.

### RNF213 activation proceeds in two steps tuned by diverse inputs

Having established ATP as the trigger of dimer opening, we next asked how additional cellular signals tune this transition, and whether opening alone is sufficient to activate the E3 ligase. We first reasoned that ubiquitination factors accessing the E3 active site should stabilize the open state by sterically preventing closure. As predicted by AlphaFold3^27^, ubiquitin binding to the RZ domain is compatible with the open but not the closed state (**Figure S5A**). Consistently, ubiquitin alone did not perturb the closed dimer but facilitated ATPγS-induced opening when present together with nucleotide (**Figure S5B**). The E2 enzyme UbcH7 showed the same behaviour: it did not alter dimer distribution on its own but potently retained the open state in the presence of ATPγS (**Figure S5C**). Thus, ATP-dependent opening creates a temporal window during which ubiquitination factors can bind and, once engaged, reinforce the open configuration.

The ability of ubiquitin and UbcH7 to stabilize the open state suggested that protein modifications affecting the core interface may themselves influence the open-closed equilibrium. To this end, RNF213 exhibits a remarkably pronounced auto-ubiquitination activity (**Figure S3A**). When ATPγS-primed RNF213 was incubated with ubiquitin-loaded E2∼Ub, we observed progressive auto-ubiquitination concurrent with a near-complete shift of the closed dimer into the open state that persisted for extended periods (**Figure 3A-B**). These data support a feed-forward mechanism in which auto-ubiquitination opposes reformation of the closed dimer and traps RNF213 in an activation-competent configuration.

**Figure 3:**
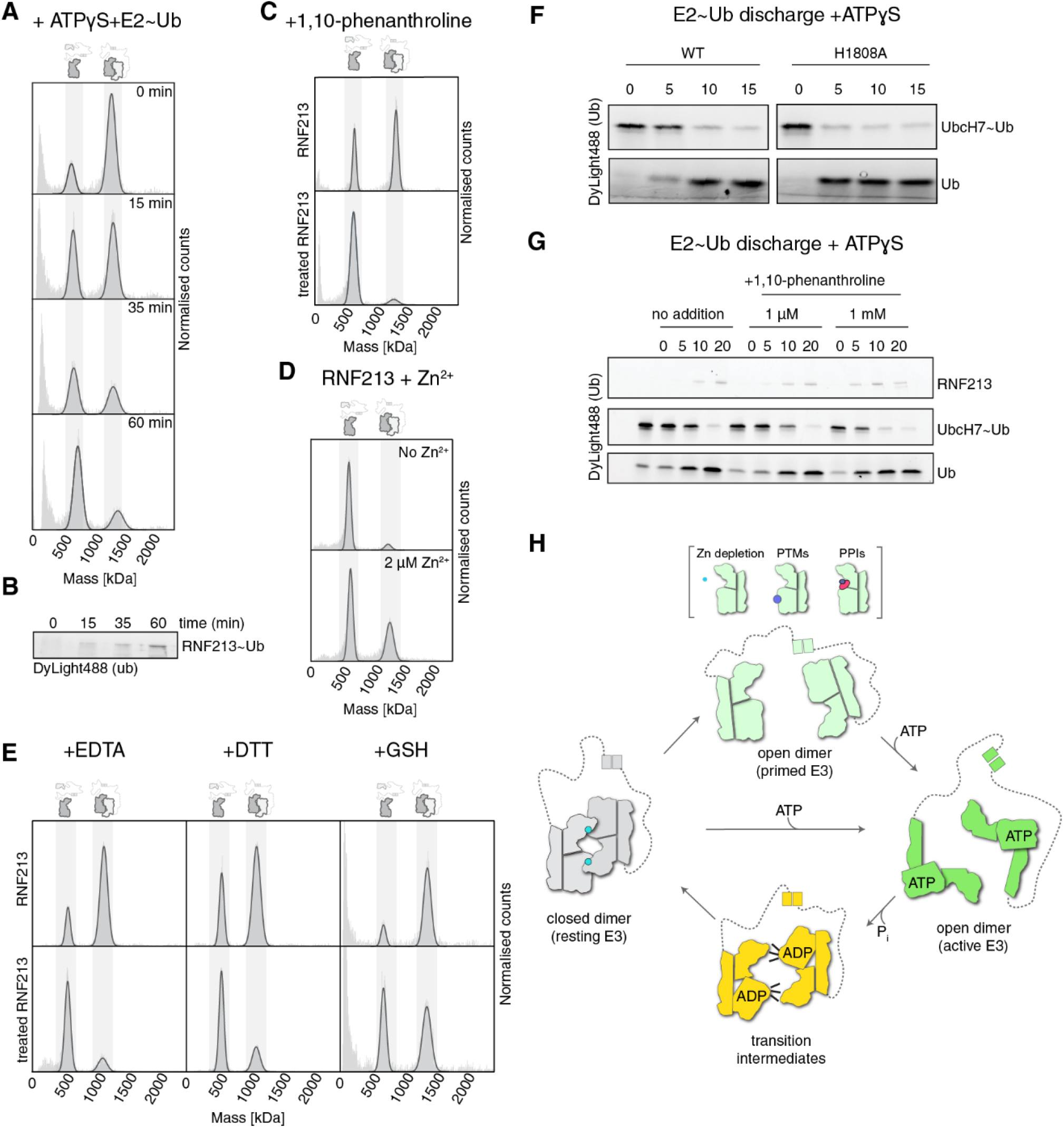
RNF213 activation proceeds in two steps tuned by diverse inputs. **(A)** Mass photometry of RNF213 + ATPγS + E2∼Ub time course. **(B)** SDS-PAGE analysis of indicated mass photometry samples in (**Fig. 3A**) showing RNF213 auto-ubiquitination. **(C)** Mass photometry of RNF213 in the absence (top) and presence (bottom) of the Zn^2+^ chelator 1,10-phenanthroline. **(D)** Mass photometry of the open dimer (LMW, **Fig. 1A**) upon titration of ZnCl_2_. **(E)** Mass photometry of the closed dimer (HMW, **Fig. 1A**) in the absence (top) or presence (bottom) of EDTA, DTT or GSH. **(F)** E2∼Ub discharge assay of ATPγS-treated RNF213 WT vs. H1808A. **(G)** E2∼Ub discharge assay of RNF213 + ATPγS in the absence or presence of 1,10-phenanthroline. **(H)** Model of how ATP binding and hydrolysis enables RNF213 to cycle between resting (closed dimer) and activated (open dimer) states in a tightly controlled manner.

The identification of Zn^2+^ coordination at the core interface pointed to a distinct mechanism that may regulate the open-closed equilibrium. Consistently, chelation of Zn^2+^ with 1,10-phenanthroline destabilized the closed dimer, biasing RNF213 toward the open state (**Figure 3C**). This effect was reversible, as addition of Zn^2+^ promoted re-formation of the closed assembly in a concentration dependent manner (**Figure 3D**). Other perturbations predicted to impair Zn^2+^ coordination (EDTA, imidazole) or to alter thiol chemistry (reducing agents DTT and GSH) produced similar shifts (**Figure 3E, Figure S5D**). Thus, Zn^2+^ coordination is a major determinant of the open-closed equilibrium, suggesting that RNF213 architecture is responsive to Zn^2+^ availability and cellular redox conditions.

We finally explored the relevance of the open-closed transition for E3 activity. Although disruption of the Zn^2+^-stabilized core interface favoured formation of the open dimer, the H1808A mutant did not exhibit increased basal E2∼Ub discharge activity (**Figure S5E**), indicating that opening alone is insufficient for E3 activation. However, the H1808A mutation stimulated ATPγS-dependent E2∼Ub discharge to levels exceeding those of wild-type RNF213 (**Figure 3F**). Likewise, treatment with 1,10-phenanthroline enhanced the rate of ATP-dependent E3 activity of the wild-type protein (**Figure 3G**), reflecting a mild stimulatory effect and the fine-regulation of RNF213 via its Zn^2+^ coordination site.

Collectively, these findings indicate that RNF213 operates through a two-step mechanism. First, ATP binding shifts the equilibrium from a closed to an open dimer, exposing the buried catalytic machinery. While this initial transition is modulated by Zn^2+^ coordination, redox state, ubiquitination factors and post-translational modifications, opening itself is not sufficient for E3 activity stimulation. ATP-dependent remodelling of the AAA module and propagation of these conformational changes through the N-arm and E3 modules drives a second transition that converts this primed dimer into a fully functional E3 ligase. ATP therefore serves a dual role as both the mechanical trigger for dimer opening and the direct driver of E3 activity, while external inputs can lower the threshold for activation by modulating the open-closed equilibrium **(Figure 3H)**.

### Moyamoya mutations destabilize the autoinhibited RNF213 dimer

Moyamoya angiopathy (MMA) is strongly associated with missense mutations in RNF213, many of which cluster within the E3 module^7,8,10,29,30^. Although disease variants have been linked to altered ubiquitination activity and inflammatory signalling^31^, the molecular basis of their pathogenic effects has remained unclear. Mapping MMA variants with the highest CADD scores, a predictor of variant deleteriousness (**Table S2**), onto the closed RNF213 dimer revealed that many localize near the core-core interface (**Figure 4A**), raising the possibility that disease mutations activate RNF213 by destabilizing the closed, autoinhibited dimer. Notably, several variants occur in regions adjacent to, rather than directly within, the dimer interface. Among these are highly pathogenic substitutions in the RING domain^31^ and multiple variants affecting internal regions of the E3 module. To better understand the distribution of the most severe mutations, we screened for de novo RNF213 variants in patients with early-onset MMA and identified a cluster of mutations proximal to the RING domain, defining a discrete disease hotspot at the interface of all E3 subdomains (**Figure S6E**; **Table S3**).

**Figure 4:**
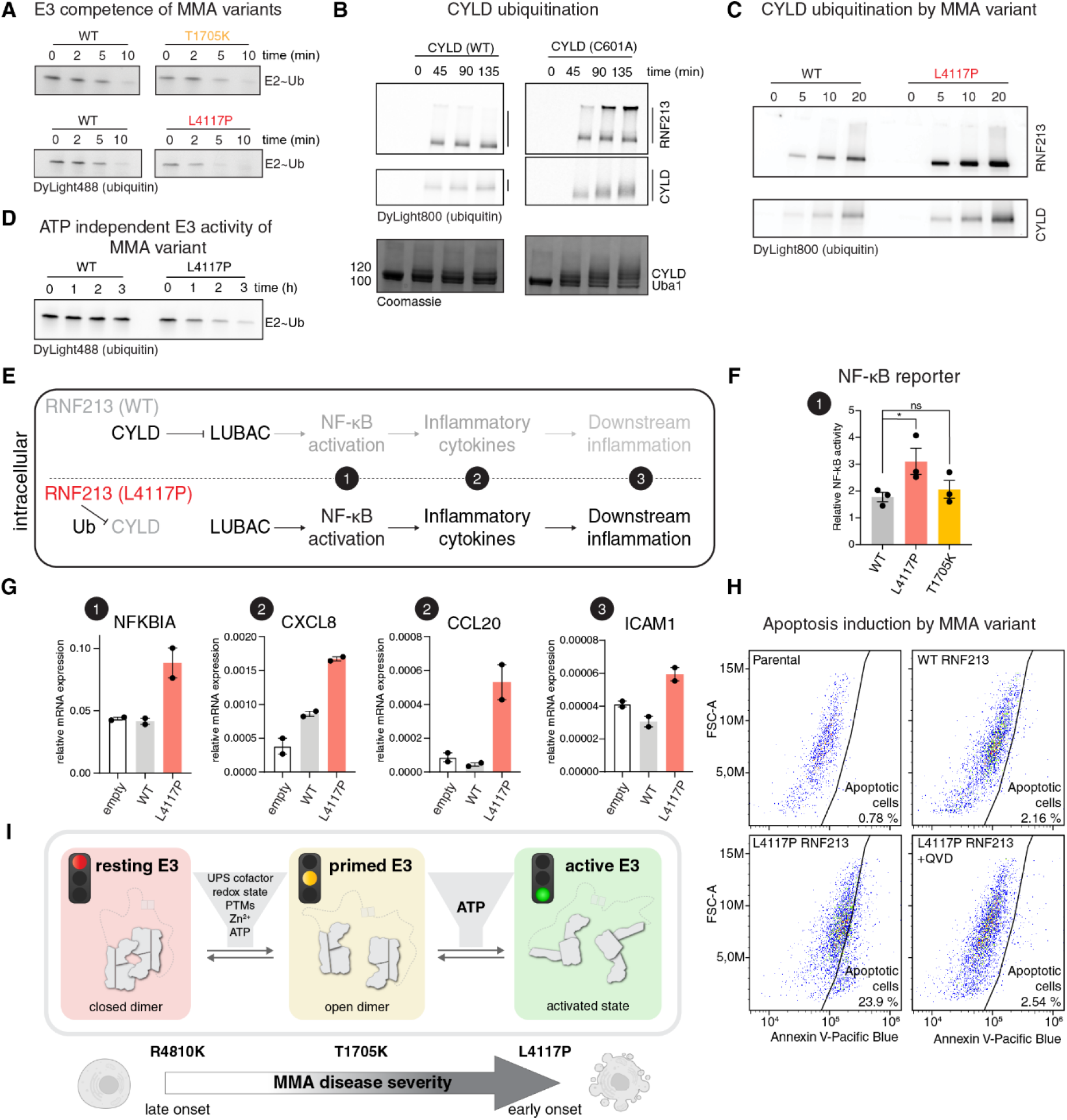
Moyamoya angiopathy mutants destabilize the autoinhibited RNF213 dimer. **(A)** Dimer model with MMA mutations (**Table S2**) mapped on protomer A. De novo, early onset MMA variants are shown in red, and other disease variants with a CADD score of >15 in gold. Residue L4117, identified as a recurrent site of MMA mutation in this study, is highlighted in magenta. **(B)** 2D cryo-EM class averages of the L4117P mutant showing single protomers of open dimers, with diffuse E3 modules and detached N-arms (labelled). **(C)** Cryo-EM models and maps (all lowpass filtered to 5 Å) of WT protomer A (this study), hsRNF213 monomers PDB 8s24 and 9jw1, and L4117P in grey, with residues 4103-4120 displayed in green to highlight ordered density for this loop (green) in all WT structures that is absent in L4117P cryo-EM analysis. **(D)** Left: Protomer A of apo dimer shown as cartoon (dark grey) and contour (light grey) to illustrate positioning of E3 module relative to AAA module. Right: Cryo-EM model of L4117P (red) overlaid with the WT contour (grey), highlighting an ∼8 Å shift of the E3 module in L4117P relative to WT RNF213, with the AAA modules aligned. Dimer interface is highlighted with observed clash in the case of L4117P. Insets 1 and 2 (shown in **Fig. 4E** and **4F** respectively) are indicated. **(E)** Zoomed view of E3-back region at the dimer interface, highlighting its disruption in L4117P, with Cys3854 – a residue of the shared Zn^2+^ binding site – highlighted in cyan. Rotation relative to the view in **Fig. 4D** is indicated in the icon. **(F)** Zoomed view illustrating the shift of the E3 module in L4117P (red) relative to WT (grey), disrupting its interaction with the N-arm. Rotation relative to the view in **Fig. 4D** is indicated in the icon. **(G)** Mass photometry of R4810K, T1705K and L4117P, showing the graded effects of mild, moderate and severe MMA mutations on shifting the open-closed transition. **(H)** Mass photometry of the L4117P upon titration of ZnCl_2_ showing partial closed dimer formation can be promoted in this variant.

To determine how mutations at this distinct hotspot affect RNF213 architecture and function, we focused on L4117P, identified independently in two children presented with recurrent ischaemic strokes, moyamoya angiopathy and systemic manifestations including pulmonary arterial hypertension and liver cytolysis (**Supplementary Information**) and showing severe abnormalities in cerebral and pulmonary circulation (**Figure S6A-C**). Analysis of the recombinant L4117P protein by cryo-EM showed that this MMA variant almost exclusively adopted the open dimer configuration, with many particles displaying an outward-swung N-arm characteristic of the fully activated state (**Figure 4B**). Although the increased flexibility of the E3 module hindered high-resolution cryo-EM reconstruction, we were able to determine a structure of L4117P with the N-arm loosely aligned to the protein core (**Figure S7**). The L4117P substitution disrupted a highly conserved loop - and MMA mutation hotspot - within the E3 module (residues 4103-4120) (**Figure S6D**) that is otherwise well ordered in all available wild-type structures (**Figure 4C**). This local perturbation propagated across the E3 module, rearranging its sub-domains and reorienting the module by ∼8–10 Å relative to the AAA unit (**Figure 4D**; **Figure S6F**). This disrupted both the dimer interface and the intramolecular docking site of the N-arm (**Figure 4E-F**; **Figure S6G**). These data directly show how a mutation distal to the dimer interface can allosterically remodel the E3 module and transform the autoinhibited dimer into the activated state.

Among MMA-associated variants, Leu4117 represents a particularly informative allelic series as different substitutions are associated with different clinical outcomes: L4117P and L4117R, identified here, are associated with severe early-onset disease, whereas the substitution L4117I is linked to later-onset manifestations, likely due to the conservative nature of this mutation^7^. This variability prompted us to ask whether other MMA variants with differential clinical severity exert graded effects on the open–closed equilibrium. As representative examples spanning the clinical spectrum, we examined the founder mutation R4810K, associated with incomplete penetrance and a low CADD score (7.4), T1705K located near the Zn^2+^-dependent interface (CADD 23.5), and the severe de novo variant L4117P (CADD 28). Mass photometry revealed increasing destabilization of the closed dimer across this series (**Figure 4G**). R4810K was mostly present in the closed dimeric form, similar to wild-type RNF213, T1705K produced a mixed population of open and closed dimers, and L4117P was found almost exclusively in the open state. The severity of disease therefore scaled with the degree to which the closed dimer was destabilized. Strikingly, raising Zn^2+^ partially restored the closed dimer of L4117P (**Figure 4H**), even though this residue lies far from the Zn^2+^ site. Despite its severity, the variant thus remains sensitive to the Zn^2+^-stabilized interface that controls the dimeric switch in wild-type RNF213, pointing to a possible route for therapeutic intervention. Together, these data show that diverse MMA variants can act through one common route, destabilizing the closed dimer to a graded extent which tracks with CADD score.

### Severe MMA variants bypass ATP control and enhance inflammatory signalling

To determine how destabilization of the autoinhibited RNF213 dimer affects downstream function, we compared E3 activity, substrate ubiquitination, NF-κB signalling and resulting cellular phenotypes across the representative MMA-associated alleles. Consistent with the mass photometry measurements, the severe variant L4117P displayed markedly increased E2∼Ub discharge activity, whereas T1705K exerted only a modest activating effect (**Figure 5A**). To determine whether these differences in basal E3 activity affect substrate ubiquitination, we reconstituted the modification of the K63-specific deubiquitinase CYLD (**Figure 5B**). In accordance with previous studies implicating RNF213 in inflammatory signalling^11,32^, inactive CYLD accumulated substantially more ubiquitin than the active deubiquitinase showing that RNF213 predominantly assembles K63-linked ubiquitin chains. Notably, the severe MMA mutant L4117P enhanced CYLD ubiquitination, whereas the moderate variant T1705K displayed activity comparable to wild-type RNF213 (**Figure 5C**; **Figure S6H**). Thus, destabilization of the restrained dimer state is reflected not only in increased intrinsic E3 activity, but also in enhanced ubiquitination of a biologically relevant substrate.

**Figure 5:**
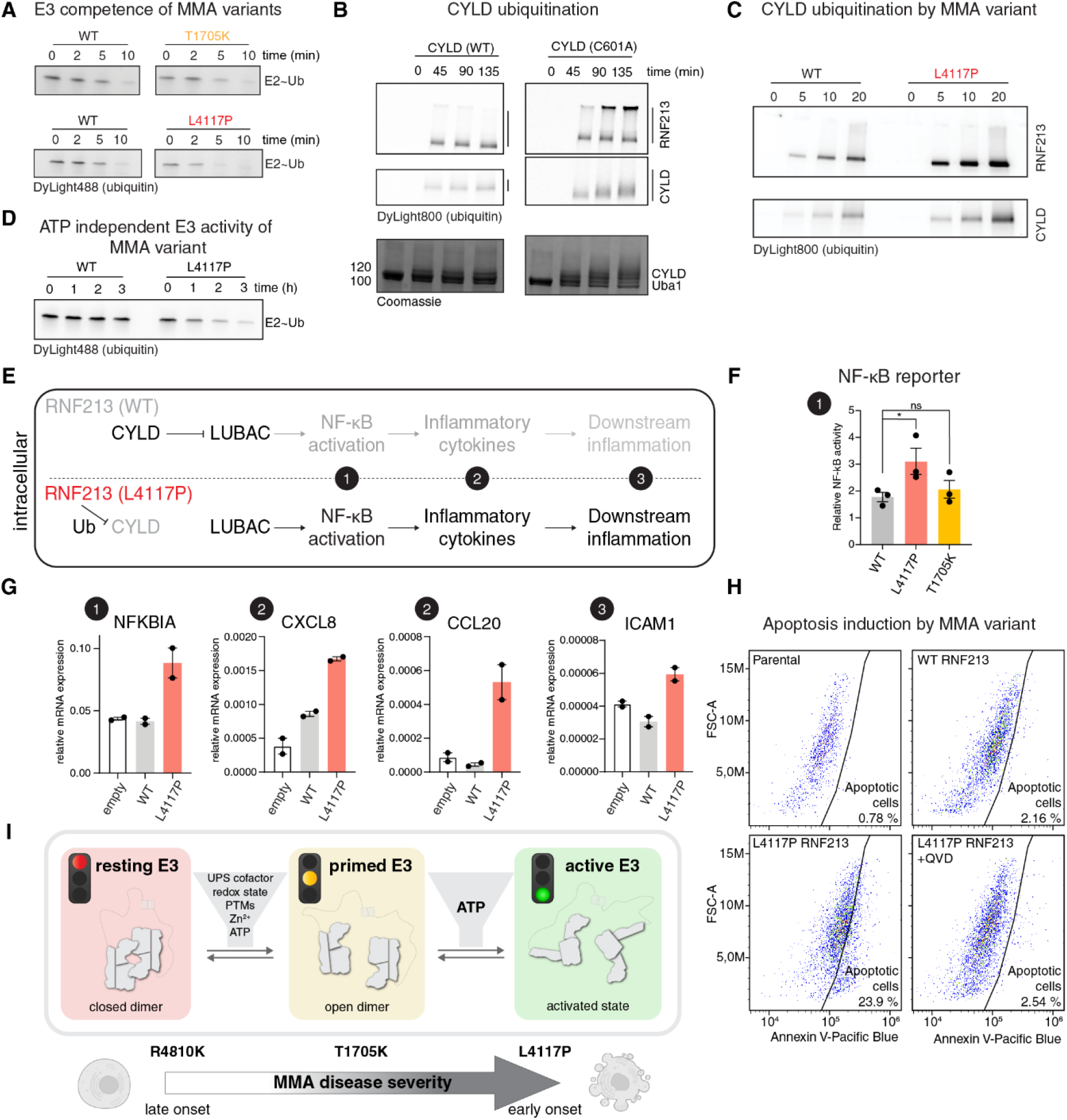
Severe Moyamoya mutations enhance RNF213 E3 activity, inflammatory signalling and apoptosis. **(A)** E2∼Ub discharge assays for WT RNF213 vs T1705K (moderate) and L4117P (severe) MMA variants. **(B)** Ubiquitination of WT or C601A (inactive) CYLD by WT RNF213. Auto-ubiquitination and CYLD ubiquitination shown in DyLight800 channel (labelled ubiquitin). Coomassie staining below shows poly-ubiquitination of CYLD. **(C)** Ubiquitination of inactive CYLD by WT or L4117P RNF213 using labelled ubiquitin. **(D)** E2∼Ub discharge assays for WT RNF213 vs L4117P in the absence of nucleotide stimuli, showing ATP-independent activity of L4117P. **(E)** Schematic illustrating proposed mechanism of RNF213 mediated induction of NF-κB signalling by ubiquitination of CYLD, allowing LUBAC-mediated signalling and subsequent inflammatory cytokine upregulation and downstream effects. Numbers 1, 2 and 3 represent steps in the pathway that are probed in subsequent figures. **(F)** Induction of a model NF-κB promoter by overexpression of RNF213 variants. T-REx-293^RNF213 KO^ cells co-transfected with empty vector/mCherry-tagged RNF213 WT, L4117P or T1705K variants, NF-κB-Firefly luciferase (FLuc) and thymidine kinase promoter-Renilla luciferase (RLuc). FLuc/RLuc ratios for each RNF213 variant compared to empty vector, shown on y-axis as Relative NF-κB activity. Data points are means from n = 3, error bars are S.E.M. Adjusted P values from a one-way ANOVA test are: WT (P = 0.2450), L4117P (P = 0.0033) and T1705K (P = 0.0950). **(G)** Bar charts for RT-qPCR measuring relative mRNA abundance of highlighted factors, with number icons indicating how each factor fits into model **(E). (H)** FACS analysis of HEK293T cells, either parental, or with Doxycycline-inducible expression of mCherry-WT RNF213 or -L4117P RNF213. X-axis shows Annexin V-Pacific Blue staining which specifically stains cells in early apoptosis, y-axis (FSC-A) depicts cell size. Manual gating based on Annexin V-negative controls was conducted in FlowJo, following gating for all cells and live cells as described in **Figure S8. (I)** Model of the two-step, hierarchical activation of RNF213 by distinct stressors and the graded disruption of this mechanism by Moyamoya-associated variants, with the most severe variants bypassing ATP-dependent control. Cell pictograms created in Biorender.com.

The increased E3 activity and the formation of activated protomers in the absence of nucleotide raised the question of whether the severe L4117P variant still depends on ATP-driven activation. Whereas ATPγS was required to stimulate wild-type RNF213 and the moderate MMA variants R4810K and T1705K, L4117P retained robust E3 activity even in the absence of nucleotide (**Figure 5D; Figure S6I**). This indicates that the severe MMA mutation can bypass the normal ATP requirement for RNF213 activation. These findings suggest that extensive remodelling of the single protomer, as reflected by the detached N-arm configuration and flexible E3 module in L4117P, is sufficient to shift RNF213 towards the activated E3 state.

We next asked whether the graded biochemical dysregulation of MMA variants was reflected in cellular inflammatory signalling (**Figure 5E)**. We first expressed RNF213 variants transiently in RNF213 knockout T-REx-293 cells and monitored NF-κB activity using an NF-κB-dependent luciferase reporter. In line with the in vitro activation profile, L4117P induced strong NF-κB activation, whereas T1705K did not elicit a significant response above WT RNF213 (**Figure 5F**). Thus, the severe MMA variant, but not the moderate variant, was sufficient to activate NF-κB signalling in unstimulated cells.

To investigate transcriptional consequences of this hyperactive MMA variant, we generated stable HEK293T cell lines expressing mCherry-tagged RNF213 WT or L4117P. RT-qPCR analysis showed increased expression of the direct NF-κB target gene NFKBIA in L4117P-expressing cells compared with WT RNF213, consistent with activation of the canonical NF-κB pathway. mRNA levels of the inflammatory genes CCL20 and CXCL8 were also elevated, as was ICAM1, an adhesion molecule induced by inflammatory signalling and associated with Moyamoya disease severity^33,34^ (**Figure 5G**). These data show that expression of RNF213 L4117P is sufficient to induce a pro-inflammatory transcriptional state in the absence of an external stimulus.

During the generation and analysis of these stable cell lines, we noticed that L4117P-expressing HEK293T cells grew more slowly and that detached cells accumulated in the culture medium, suggesting reduced viability. To test whether this reflected apoptotic cell death, we performed flow cytometry analysis with Annexin V staining. Doxycycline-induced expression of WT RNF213 caused only a slight increase in apoptotic cells relative to doxycycline-treated parental HEK293T controls. By contrast, expression of RNF213 L4117P increased the apoptotic cell population by more than 20%. This effect was largely suppressed by the pan-caspase inhibitor QVD, reducing Annexin V staining to levels observed for WT and parental control cells (**Figure 5H**; **Figure S9**). Thus, the severe L4117P variant not only activates inflammatory signalling but also promotes caspase-dependent apoptosis.

Taken together, these findings show that MMA mutations destabilize the autoinhibited RNF213 dimer and shift the conformational equilibrium toward activation-competent states. The extent of this activation bias correlates with disease severity across the variants examined, yielding graded increases in intrinsic E3 activity, substrate ubiquitination and NF-κB signalling. In the most severe variant tested, structural remodelling uncouples E3 activation from ATP stimulation and is sufficient to drive inflammatory gene expression and caspase-dependent apoptosis in cells. These data provide a structural and mechanistic explanation for the strong biochemical and cellular phenotypes of severe RNF213 MMA variants.

## DISCUSSION

RNF213 is an immune-associated E3 ligase whose activity must be tightly restrained to prevent inappropriate inflammatory signalling. Here we show that RNF213 is controlled by a reversible dimeric switch. Contrary to the prevailing view of RNF213 as a monomer, our data show that it functions as a constitutive dimer that interconverts between closed, autoinhibited and open, activation-competent states. In this arrangement, E3 activity is not only set by local rearrangements within each protomer, but also by changes in how the two protomers engage one another.

Mechanistically, the dimeric switch is implemented through two distinct interfaces. A high-affinity interaction between the N-terminal ZF1 domains tethers two RNF213 protomers, whereas a second regulated interface aligns the AAA and E3 regions into a compact closed assembly. Engagement of this second interface sterically occludes the catalytic machinery, while its disruption exposes the E2-binding CTD and catalytic RZ domain. This mode of regulation, occurring at the level of oligomer re-assembly, differs from the local autoinhibitory mechanisms typical of many ubiquitin ligases^35,36^. Instead, RNF213 resembles large AAA+ enzymes such as dynein and midasin, in which nucleotide-dependent conformational changes induce a large-scale remodelling of the molecular machines to release their restrained states and permit engagement with partner proteins or substrates^37-39^.

Our findings reveal how the RNF213 dimeric switch integrates diverse biological inputs through a two-step activation mechanism. ATP, whose cellular levels rise during infection^1^, acts as the principal effector: it first opens the closed dimer and then, in the second step, remodels the separated protein cores into the fully active conformation, marked by N-arm detachment and increased flexibility of the E3 module. Additional inputs can tune the first step by shifting the open– closed equilibrium: Zn^2+^ coordination favours the restrained state, whereas reducing conditions, partner proteins such as UbcH7 and auto-ubiquitination favour the open, pre-activated state ^6,16^. In each of these cases however, the second step of forming the active E3 state remains ATP-dependent. The tuneable dimeric switch is likely subject to further regulatory inputs. For example, stress-induced phosphorylation of Y1275^11^ maps to the linker region that rearranges during dimer opening and may thus directly alter the open-closed transition. As ZF1 tethering retains RNF213 protomers in proximity, dimers can rapidly re-close upon ATP hydrolysis to prevent hyperactivation. Thus, RNF213 activation is hierarchically organized: stress- and pathogen-linked inputs tune dimer opening, whereas ATP transiently drives the open dimer into the fully active E3 state.

This safeguard mechanism also reveals the molecular basis for RNF213-associated Moyamoya disease. Rather than directly activating the E3 ligase, MMA mutations bias the dimeric switch toward activation-competent states by destabilizing the autoinhibited assembly, as directly visualized for the severe L4117P mutant. The degree of destabilization correlates with disease severity, producing graded dysregulation of E3 activity and inflammatory signalling. While mild variants lower the activation threshold imposed by the dimeric switch, severe mutations such as L4117P also bypass the second activation step, namely ATP-dependent remodelling of the E3 module, and thereby uncouple RNF213 from its normal regulation mechanism (**Fig. 5I**).

Importantly, the severe L4117P variant converted RNF213 activation into a disease-relevant cellular phenotype. In unstimulated cells, L4117P activated canonical NF-κB signalling, induced inflammatory genes including CCL20, CXCL8 and ICAM1, and triggered caspase-dependent apoptosis. This pro-inflammatory and pro-apoptotic state provides a direct cellular correlate for the biochemical hyperactivation observed *in vitro*. It also links disruption of the RNF213 dimeric switch to processes that are central to vascular disease, including inflammatory activation, altered cell adhesion and loss of cell viability. The endogenous ubiquitination events that connect RNF213 activation to these outputs remain to be resolved. Our reconstitution experiments confirm CYLD as a direct substrate linking RNF213 to NF-κB signalling. Since L4117P enhanced CYLD ubiquitination, this event could act in cells to release the brake on the LUBAC-dependent NF-κB pathway. Further potential RNF213 substrates, such as TRAF2^40^ or TRAF3^11,41^, may also contribute to cellular signalling in a context-dependent manner. We propose that the dimeric switch acts upstream of such events as a safety mechanism: when intact, it restrains RNF213 E3 activity within these immune pathways, but when destabilized by severe Moyamoya mutations, it permits constitutive inflammatory and pro-apoptotic signalling.

Strikingly, elevated Zn^2+^ concentrations can restore the closed dimer in the severe MMA variant *in vitro*, implying that disease-induced destabilization of the E3 assembly can be counteracted and identifying the dimeric switch as a potential therapeutic target.

Because this switch integrates multiple cellular inputs, it also offers an explanation for the variable penetrance of MMA-associated alleles: the risk of developing disease symptoms rises as mutations lower the activation threshold, allowing infection-associated signals including elevated ATP levels or RNF213 modifications to trigger excessive inflammatory ubiquitin signalling. This is in line with the “second-hit” model previously described for mild MMA mutation carriers, who often live symptom-free into adulthood until a secondary infection triggers disease onset^42,43^. The RNF213 dimeric switch therefore links genetic predisposition, environmental triggers and infection in Moyamoya disease and related inflammatory disease states.

## Data Availability

All data produced in the present work are contained in the manuscript

## ACKNOWLEDGEMENTS

We would like to thank the EM facility of the Vienna BioCenter Core Facilities (VBCF), particularly Harald Kotisch for collecting the cryo-EM datasets. We thank the VBCF Protein Technologies and Proteomics facilities, as well as Mathias Madalinski from Peptide Synthesis. We thank all members of the Clausen lab for their support and discussions. We would additionally like to thank Maximilian F.D. Spicer and Daniel W. Gerlich, IMBA at the Vienna Biocenter for their advice and reagents when setting up the apoptosis FACS experiment. Finally, we are grateful to Sylvie Nguyen The Tich (Hôpital Roger Salengro, Lille) for neurological evaluation of the child with the L4117P variant.

## FUNDING

This work was supported by the Leducq Foundation Network of Excellence Program grant ReVAMP (LFCR grant 21CVD03, E.T.L.) and the French priority research program on rare diseases “Programme Prioritaire de Recherche Maladies Rares du Programme Français d’Investissement d’Avenir” (E.T.L.). This work was also supported by the Austrian Research Promotion Agency (Austrian Life Science grant FO999902549), a BI Fonds fellowship (L.E.B.), Marie Skłodowska-Curie grant 847548 (D.B.G.), WWTF grants 10.47379/LS21029 (L.E.B) and 10.47379/LS21009 (L.D), a UKRI Future Leaders Fellowship MR/T043482/1 (H.S. and A.J.F.) and the European Research Council under the European Union GA No 101141607 MUSCLE-QC (T.C.). The IMP is supported by Boehringer Ingelheim. For the purpose of Open Access, the author has applied a CC BY public copyright licence to any Author Accepted Manuscript (AAM) version arising from this submission.

## AUTHOR CONTRIBUTIONS

T.C., A.F., L.E.B and V.F. designed experiments. L.E.B, V.F. M.H.W. and A.M. performed cloning, and L.E.B and L.D. performed expression and purification. L.E.B and V.F. performed biochemical experiments. L.E.B and D.B.G. generated and analysed cryo-EM data. T.L.W. performed and analysed native mass spectrometry. L.E.B, E.K. and H.S. performed the cellular assays. M.K., L.L., F.G., F.R., C.A., E.T.L., provided clinical and molecular genetics data. A.F. and T.C. supervised research and provided scientific input. T.C. coordinated the research project. L.E.B and T.C prepared the manuscript with input from all authors.

## DECLARATION OF INTERESTS

The authors declare no competing interests.

## DATA AND MATERIALS AVAILABILITY

Cryo-EM maps and atomic coordinates have been deposited in the Protein Data Bank (PDB) with accession codes 34QV (apo dimer); 29WC pdb_000029WC (int1); 29VL pdb_000029VL (int2); 29VA pdb_000029va (L4117P MMA mutant) and the Electron Microscopy Data Bank (EMDB) with accession codes EMD-100123 (apo dimer composite map), EMD-100087 (apo dimer consensus map), EMD-100089 (apo dimer E3 local refinement), EMD-100115 (apo dimer N-arm local refinement), EMD-100122 (apo dimer ATPase local refinement), EMD-100114 (apo dimer interface local refinement), EMD-100116 (apo dimer CTD local refinement), EMD-100117 (apo dimer CBM local refinement); EMD-57400 (int1); EMD-57398 (int2); EMD-57388 (L4117P).

## ETHICAL CONSIDERATIONS

The 3 affected adult patients carrying the L4117I variant and both parents of the affected child carrying the de novo L4117P variant provided their written informed consent for genetic analysis in accordance with French ethical recommendations for genetic testing. All patients for which data has been included in this study have been anonymised.

## SUPPLEMENTARY INFORMATION

Clinical summary of patient with L4117P

The patient had unremarkable prenatal and neonatal history, with normal early development. Between 0-5 years of age, they presented with paroxysmal right-sided abnormal movements and right hemiparesis triggered by crying. Brain MRI revealed an ischemic stroke in the territory of left middle cerebral artery (MCA) associated with Moyamoya disease. Despite aspirin therapy, follow-up MRI showed progression of ischemic lesions, leading to cerebral revascularization surgery by multiple burr-hole synangiosis on day 10 (**Figure S6A**). Initial fundoscopy, transthoracic echocardiography, and renal artery Doppler ultrasound were normal. Following a minor head injury, they developed contralateral (left-sided) hemiparesis and repeated focal seizures. Neuroimaging revealed a right subdural hematoma and recent ischemic lesions in the right MCA territory (**Figure S6B**). Neurological outcome was characterized by mild cerebral palsy with persistent minimal left hemiparesis. Independent walking was acquired, with language delay and a heterogeneous but globally normal neuropsychological profile at a later stage.

Still between 0-5 years of age, the onset of dyspnea led to the diagnosis of severe pulmonary arterial hypertension with right ventricular hypertrophy. The patient underwent cardiac catheterization. Pressure records were as follows: right ventricle 80/0 mm Hg, main pulmonary artery 89/19/mean 47 mm Hg, left proximal pulmonary artery 74/17/mean 41 mm Hg, right proximal pulmonary artery 87/18/mean 43 mm Hg, and mean right pulmonary capillary wedge pressure of 10 mm Hg. Cardiac catheterization revealed markedly elevated mean pulmonary arterial pressure of 47 mm Hg, with a normal pulmonary capillary wedge pressure of 10 mmHg. On selective pulmonary angiography, both proximal right and left pulmonary arteries were well developed, contrasting with severe bilateral hypoplasia of the distal pulmonary vascular tree (**Figure S6C**). The patient died between 5-10 years of age from Group A streptococcal toxic shock syndrome complicating pleuropneumonia, with refractory circulatory failure despite maximal intensive care, including extracorporeal membrane oxygenation.

## MATERIALS AND METHODS

### Cloning and expression

For insect and mammalian cell expression, RNF213 gene from H. sapiens was codon-optimized for mammalian cell expression and synthesized in fragments flanked with BsaI sites. RNF213 was synthesized with a C-terminal Twin-Strep tag. The gene was assembled via scarless GoldenGate reactions from 3 gBlocks into either a pGBdest vector (insect cells) or pCMV vector (mammalian cell expression) where the final assembled gene was flanked by a polyhedrin promoter and an SV40 polyA signal. In addition, pCMV constructs were assembled to contain an N-terminal mCherry and 5’ intron. Mutations were generated by Gibson assembly into the respective gBlock-containing plasmid. The sequence of all generated insect and mammalian cell constructs was confirmed by whole plasmid sequencing. For insect cell expression, following Golden Gate assembly, plasmids were transformed into DH10EmBacY cells and blue-white screening was used to select colonies with bacmids. Bacmids were extracted by alkaline lysis and isopropanol precipitation and then transfected using PEI into Sf9 cells for viral amplification. Protein expression was performed in Trichoplusia ni High-Five cells. Cells were infected with virus at a density of 1.25 × 10^6^ ml^-1^ and grown for three days at 27 ^o^C. Cells were harvested by 20 min centrifugation at 600 g.

### Protein purification

Insect cell expression pellets were thawed in a water bath and resuspended at a 1:2 ratio in lysis buffer (25mM HEPES, 500 mM KCl, 0.25 mM TCEP, pH 7.2) with 1 tablet of cOmplete EDTA-free Protease Inhibitor. Cells were lysed using a tissue homogenizing douncer. For DNA precipitation, 5 (w/v) % PEI solution was added dropwise while stirring to a final concentration of 0.05 (w/v) % and incubated at 4 °C for 15 minutes. Lysis clarification was performed at 18000 rpm for 45 minutes using a fixed-angle rotor centrifuge at 4v°C. For His-tagged constructs, the supernatant was filtered by using a 0.45 µm filter and applied onto a 5mL HisTrap column pre-equilibrated in lysis buffer. The column was washed for 75 mL with 25mM HEPES, 1 M KCl, 0.25 mM TCEP, pH 7.2 followed by a 40 mM imidazole wash for 100 mL. The protein was eluted by a linear gradient ranging from 10-375 mM imidazole for 75 mL. For Strep-tagged constructs, the supernatant was filtered by using a 0.45 µm filter and applied onto a 5 mL StrepTrap XT column pre-equilibrated in lysis buffer. The column was washed with 75 mL of lysis buffer before step elution with 1X BXT (iba Life Sciences), diluted from 10X in MilliQ water. For both Strep- and His-tagged constructs, eluted fractions were analysed by SDS PAGE and pooled accordingly. From this point, both Strep- and His-tagged constructs were treated identically. The pooled sample was diluted to a final conductivity of ∼15 mS/cm in 25mM HEPES, 0.25 mM TCEP, pH 7.2 and loaded onto a 5 mL Resource Q anion exchange column pre-equilibrated in 25mM HEPES, 100 mM KCl, 0.25 mM TCEP, pH 7.2. The column was washed in the same buffer before protein elution with a linear gradient of increasing KCl over 20 CV to 1 M. Eluted fractions were analysed by SDS PAGE and concentrated. Final sample was injected onto a Superose 6 10/300 GL column equilibrated in 25mM HEPES, 200 mM KCl, 1 mM TCEP, pH 7.2. Eluted RNF213 fractions were pooled according to SDS PAGE, concentrated to >1 mg/mL, aliquoted and flash-frozen in liquid nitrogen and stored at –70 °C. The three-step purification procedure was performed using a ÄKTA pure chromatography system at 4 °C. To determine the final protein concentrations absorbance at 280 nm was measured (extinction coefficient: 582710 M^-1^ cm^-1^).

### Cryo-EM grid preparation and data collection

For grid preparation, 0.6 mg/mL hsRNF213 in 25 mM HEPES pH 7.2, 200 mM KCl, 0.25 mM TCEP was applied onto a freshly glow-discharged (180 seconds) UltrAuFoil R2/2 on 200 mesh grid. Grids were blotted for 1.2 seconds before rapid freezing in liquid ethane using a Leica GP1 plunge-freezer. A Glacios TEM (Thermo Fisher) equipped with a Falcon 3 detector was used to screen grids. For data collection, the grids were transferred to a Titan Krios G4 (Thermo Fisher) with a Falcon 4EC detector operated by the Research Institute of Molecular Pathology, Austria. Data was collected using the EPU software (Thermo Fisher) with 130,000x magnification (0.951 Å pixel size) and a total dose of 50 e/Å^2^. Patch Motion correction was performed on-the-fly using CryoSPARC Live.

### Cryo-EM data analysis

The HsRNF213 dimer cryo-EM structure was determined via collection of a single dataset. Processing was conducted using the specific strategy detailed in **Figure S2, Figure S5** and **Figure S8**. On-the-fly CryoSPARC live session was detached from the live server and transferred to CryoSPARC v4 for processing^44^. Particle picking and removal of junk particles was carried out by rounds of 2D classification. From the remaining 2D classes, templates were generated and used for template picking. Resulting picks, after junk was again removed by 2D classification, were extracted in a 512 px box size downscaled initially to 128 px. 3 Ab-initio reconstructions were generated from the remaining particles, and these volumes were used as input for several rounds of heterogeneous refinement to remove additional junk in 3D. Remaining good particles were then re-extracted with a 512 px box size downscaled only to 384 px (1.27 Å/px) (note: particles used for ATPase local refinement were re-extracted without downscaling), and the newly extracted particles were used for 3D refinement with either C1 or C2 symmetry enforced as described in the respective Supplementary Figures. For the hsRNF213 apo dimer, the particle stack was then symmetry expanded and, using local maps, 3D classified before a subset of the classes were locally refined to generate the final maps. A composite map was made in ChimeraX by aligning these focused maps onto the initial 3D-refined map with C2 symmetry, and scaling before combining using the volume maximum command.

Additional cryo-EM structures of int1, int2 and L4117P presented in this manuscript were similarly processed, with further detail in their respective Supplementary Figures. In contrast to the RNF213 apo dimer, each additional cryo-EM structure was modelled from a single map, plus a local refinement of the AAA module for the int1 and int2 states to assess nucleotide occupancy. Particles contributing to the final models of each were, as above, extracted with a 512 px box size downscaled only to 384 px (1.27 Å/px), with C1 symmetry enforced for each 3D refinement step.

### Molecular model building and refinement

For the HsRNF213 apo dimer, the 6 focused maps (E3 module, ATPase, N-arm, CBM, CTD and dimer interface) were combined as described above to create a composite map used for modelling. The human RNF213 monomer (PDB: 8s24) was used as the starting point for model building of the dimer. Where the resolution between the monomer and dimer differed, regions that had been excluded from 8s24 were first modelled using ModelAngelo^45^ then built into the existing model using Coot and Isolde^46,47^. For the CBM, AF3 was used to predict the entire domain, which was docked into the dimer model and rigid body fitted. Once all regions visible in our EM density were built, the final model was obtained by rounds of modelling in Coot and Isolde, followed by refinement using Phenix Real Space Refine^48^. Final local resolutions and model to map fitting are detailed in **Figure S2**, and refinement statistics can be found in **Table S1**.

For Int1, Int2 and L4117P cryo-EM structures, models were built from single EM density maps for the sake of illustrating a single state within the heterogenous datasets. In all cases, the hsRNF213 apo dimer, or a single protomer thereof, was used as a start point for model building. Modelling again was conducted using Coot and Isolde, and where density was absent, the corresponding region in the models were removed.

To analyse heterogeneity in RNF213 + ATPγS dataset, 3D variability analysis was conducted in cryoSPARC, with 3DVA output intermediate mode enabling output of particle stacks contributing to each intermediate frame along the single component^49^.

### Negative stain electron microscopy (EM)

Continuous carbon coated copper/palladium grids (Agar Scientific) were glow discharged at 25 mA for 60 seconds using a BAL-TEC sputter coater CD005. RNF213, diluted to 0.025 mg/mL in 25 mM HEPES pH 7.2, 200 mM KCl, 0.25 mM TCEP, was spotted on grids, incubated for one minute, blotted, washed with 4 μL 2 (w/v) % uranyl acetate, blotted again, and incubated for one minute with 4 μL 2 (w/v) % uranyl acetate before final blotting and drying. Data were collected on a 200 kV FEI Tecnai G2 20 (Thermo Fisher Scientific) at 50,000 x magnification with a 2.21 Å pixel size using SerialEM. All data processing was carried out in CryoSPARC v4 on with a 2x binned 288 px box size.

### In vitro ubiquitination assays

In vitro ubiquitination reactions were performed with 0.5 μM human UBE1, 8 μM human UbcH7, 40 μM DyLight800/488-Maleimide-Ubiquitin^23^, 5 μM BSA monomer and 0.15 μM RNF213 using 25 mM HEPES, 150 mM KCl, 10 mM MgCl_2_, and 2 mM TCEP pH 8.0 as reaction buffer. Reaction mixes were prepared on ice and initiated by addition of 1 mM ATP, incubated at 30 °C and quenched for timepoint collection by addition of SDS-PAGE sample buffer.

### E2∼Ub discharge assays

The ubiquitin assay in discharge format was performed by pre-charging UbcH7 with DyLight488 labelled ubiquitin. In short, 1.5 µM Uba1, 90 µM UbcH7, 1 mM ATP and 150 µM ubiquitin (1:150 unlabelled vs DyLight488 labelled) were mixed in buffer containing 25 mM HEPES pH 8, 150 mM KCl, 1 mM TCEP, 10 mM MgCl_2_ and incubated for 30 minutes at 37 °C. Next, the reaction was mixed with Ni Sepharose High Performance beads (Sigma-Aldrich) and incubated at 4 °C for 1 hr with rotation to remove His-tagged Uba1. After 3 X washing and centrifugation steps, the resulting flowthrough was injected onto a Superdex™ 75 Increase 10/300 GL size exclusion column pre-equilibrated in 2CV of 25 mM HEPES pH 7.2, 200 mM KCl, 0.25 mM TCEP. The eluted fractions were analysed by SDS-PAGE and fractions containing charged UbcH7∼Ub were pooled, concentrated and flash frozen at –70 °C.

In vitro E2∼Ub discharge assays were performed with 7 µM UbcH7∼Ub, 5 µM BSA, 1 mM ATPΓS (unless no nucleotide control stated) and 0.15 µM RNF213 in 25 mM HEPES, 150 mM KCl, 10 mM MgCl_2_, and 2 mM TCEP pH 8.0 as reaction buffer. Mastermixes (with UbcH7∼Ub and RNF213 in separate reactions) were made up on ice and initiated by mixing 1:1 and incubating at 30 °C with timepoints taken and quenched with the addition of non-reducing SDS-PAGE sample buffer.

### Mass Photometry

Experiments were performed using a OneMP or TwoMP mass photometer (Refeyn Ltd.) controlled by the AcquireMP application, following the standard operating procedure^26,50^. A one-minute acquisition time in the Regular field of view was used. To obtain average species masses, particle event histograms were automatically Gaussian fitted with the DiscoverMP software. Prior to each data acquisition session, the machine was calibrated by measuring NativeMark™ (Thermo) protein standard and correlating radiometric contrast with known molecular weights of the standard in DiscoverMP. Samples were first prepared in a 400 nM stock solution in 25 mM HEPES pH 7.2, 200 mM KCl, 0.25 mM TCEP and prior to measurement were diluted 10 x in room temperature PBS. Data were plotted in the DiscoverMP application. For time course experiments, samples of RNF213 (+ATPγS, **Figure 2B** and +ATPγS+UbcH7∼Ub, **Figure 3A**) were made by first making an 800 nM mastermix of RNF213 and mixing 1:1 with 2 mM ATPγS and 10 μM UbcH7∼Ub (where applicable). Upon mixing, an initial sample was taken and recorded as time 0, then samples were incubated at 30 °C and further timepoints taken at the indicated times. For the RNF213+ATPγS+UbcH7∼Ub time course, when 2 μL mass photometry samples were taken, a further 4 μL sample was taken and mixed 1:1 with non-reducing loading dye for SDS-PAGE analysis (**Figure 3B**).

### Native mass spectrometry

The ZF1 peptide was dissolved in 100 mM ammonium acetate, pH 7.4, 2 mM ZnCl_2_ and then purified by size exclusion chromatography to remove excess Zn^2+^ from the sample. Native mass spectrometry analysis was performed on a Waters, Synapt G2-Si equipped with a Waters Z-spray nano source. The peptides were infused at a concentration of approximately 40 µM and data was collected in positive ion mode. The source temperature was set to a constant 75 °C and samples were ionised at a capillary voltage of 1.2 keV. Data was collected for 1 minute and a scan was taken every 1 second. The data was combined and the spectra analysed using MassLynx v4.2.

### ZF1 Size Exclusion Chromatography

Peptides of WT ZF1 (sequence: MECPSCQHVSKEETPKFCSQCGERLPPA) or F17A ZF1 (sequence: MECPSCQHVSKEETPKACSQCGERLPPA) were obtained from VBCF Peptide Synthesis as lyophilised powder. 1 mg peptides were solubilised in 25 mM HEPES pH 7.2, 200 mM KCl, 2 mM ZnCl_2_ to 850 μM. Superdex 30 Increase 10/300 GL column (Cytiva) was pre-equilibrated in 2 CV of 25 mM HEPES, 200 mM KCl, 1 mM TCEP before 100 μL of peptide diluted to 85 μM was injected. Column was washed in 1CV buffer between each peptide run.

### Cell culture

All cells used in this manuscript, namely T-REx-293^RNF213 KO^, HEK293T, HEK293T PiggyBac TetON-mCherry-WT RNF213 and HEK293T PiggyBac TetON mCherry-L4117P RNF213 were maintained at 37 °C, 5 % CO_2_. Cell media for both maintenance and seeding, unless otherwise stated, was Dulbecco’s Modified Eagle Medium (DMEM, Gibco), 10 % FBS, 2 % L-glutamine.

HEK293T cells stably expressing doxycycline-inducible mCherry-RNF213 fusion protein were generated using the piggyBac transposition system (polyclonal). The resulting cell lines utilised in this manuscript were HEK293T (no RNF213 construct), PiggyBac TetON-mCherry-WT RNF213, and PiggyBac TetON mCherry-L4117P RNF213.

### NF-κB signalling assay

T-REx-293^RNF213 KO^ cells - generated as described in^1^ - seeded at 100,000 cells per well of a 24-well plate, were transfected the next day with 200 ng empty CMV promoter vector or mCherry-tagged RNF213 WT, L4117P or T1705K variants, alongside 20 ng pGL4.32 (luc2P/NF-κB-RE/Hygro) ^29^ (Promega) and 10 ng pRL-TK (Promega). 24 h post transfection, cells were processed using the Dual-Luciferase Reporter Assay System method (Promega, E1910). Cells were lysed in 1x Passive Lysis Buffer and 20 μL lysate mixed with 30 μL Luciferase Assay Reagent II and firefly luminescence quantified using a Glomax luminometer (Promega). The reaction was quenched with the addition of 30 μL Stop & Glo reagent, followed by sequential reading of renilla luminescence (480 nm). Firefly:renilla ratios from RNF213 transfection were compared to those derived from empty CMV promoter vector.

### RT-qPCR

#### Cell seeding, maintenance and transfection

HEK293T (no RNF213 construct), PiggyBac TetON-mCherry-WT RNF213, and PiggyBac TetON mCherry-L4117P RNF213 cells were seeded at 200,000 cells/well in 12-well plates in DMEM maintenance media. 24 hrs after seeding, parental HEK293T (no RNF213 construct) were transfected with jetOPTIMUS® DNA transfection reagent (Polyplus) by diluting 1 μg pcDNA3.1 in the provided buffer, mixing with jetOPTIMUS® reagent at a 1:2 ratio, incubating at room temperature for 10 min, then adding dropwise to cells in complete growth medium, to generate empty plasmid controls. At same time, all cells were treated with 200ng/mL Doxycycline hyclate (Sigma-Aldrich, Cat.No. D5207).

#### Reverse transcription

Cells were washed twice with PBS and lysed in 100 μL of Lysis Buffer (RNeasy Mini Kit, Qiagen). Total RNA was isolated using the RNeasy Mini Kit (Qiagen) according to the manufacturer’s protocol.

The final mixture for reverse transcription contained 1 μg/μL of the extracted RNA and 1 μg/μL Oligo (dT) 18 primer (Thermo Fisher Scientific, Cat.No. S0131) per sample. The mixture was heated for 5 min at 65°C and cooled down on the ice. Master mix of 1x Reaction buffer for RT (Thermo Fisher Scientific), 1 mM dNTPs mix (Thermo Scientific, Cat.No. RO192) and 200 U/μL RevertAid Reverse Transcriptase (Thermo Fisher Scientific, Cat.No. EP0441) per sample were added. The mixture was heated for 60 min at 42°C, followed by 70°C for 10 min.

#### Real-Time PCR

TaqMan™ Fast Advanced Master Mix for qPCR (Thermo Fisher Scientific, Cat.No. 4444556) and pre-designed RT-PCR assays were used (listed below).

Briefly, the master mix and the samples were pipetted into a 96-wells plate (Roche) in duplicates and the plate was centrifuged at 1200 g for 2 min, 4 °C. Analysis was performed in Biorad CFX 384 Real Time Cycler, with the program: 50°C/2 min; 95°C/20 sec; 95°C/3 sec; 60°C/30 sec. Steps 3-4 were repeated 40 times.

Results are expressed as the fold change of the target gene and the average of reference genes (*HPRT* (Hypoxanthine Phosphoribosyltransferase 1), RPL13A (*Large ribosomal subunit protein uL13))*, determined by the relative quantification 2^-ΔCq^ method.

#### Pre-designed RT-PCR assays

Catalogue number: 4453320

Assay ID: Hs00355476_m1

Gene Symbol: CCL20

Catalogue number: 4453320

Assay ID: Hs00174103_m1

Gene Symbol: CXCL8

Catalogue number: 4453320

Assay ID: Hs00164932_m1

Gene Symbol: ICAM1

Catalogue number: 4453320

Assay ID: Hs00355671_g1

Gene Symbol: NFKBIA

### Monitoring caspase-induced apoptosis with FACS

#### Cell maintenance and seeding

HEK293T (no RNF213 construct), PiggyBac TetON-mCherry-WT RNF213, and PiggyBac TetON mCherry-L4117P RNF213 as in RT-qPCR analysis were used for all FACS experiments and maintained as above. For FACS, 20,000 cells/well were seeded into a 96-well plate 24 hrs prior to experimentation. All seeded cells were treated with doxycycline as above to either induce mCherry-RNF213 expression, or as a control for Dox-induced apoptosis. Where applicable, cells were additionally treated with the pan-caspase inhibitor Q-VD-OPh (10 μM final concentration) at the time of seeding.

#### Cell harvesting, staining and analysis

The following day, cell media was removed into a V-bottomed 96-well FACS-compatible collection plate (Nunc), cells were washed with PBS, and wash was added to the collection plate, then trypsinised in 20 μL TrpLE™ Express no phenol red (Gibco) for 30 seconds, before cell suspension was added to the same collection plate. Plate of harvested cells, media and wash was then centrifuged for 4 min, 125 g. During centrifugation, Annexin V-Pacific Blue stain (Thermo, A3512) was prepared by 1:50 dilution in 1x PBS. After centrifugation, media was aspirated and cells were resuspended in 70 µL Annexin V-Pacific Blue-PBS, or 1x PBS only for no-stain controls.

Flow cytometry analysis was performed on an Agilent NovoCyte Penteon five laser flow cytometer, with a flow rate of 35 µL/min. Mixing was performed before acquisition of each sample. Flow cytometry data was analysed using FlowJo v10.10.0.

### LC-MS/MS and bioinformatic analysis

#### Sample preparation

10 µl of the 2 peak fractions of size exclusion chromatography (Figure 1A) contained in 25 mM HEPES, 200 mM KCl, 1 mM TCEP, pH 7.2 were mixed 1:1 with 10 M urea in 100 mM ammonium bicarbonate (ABC). Proteins were reduced with 10 mM Dithiothreitol (DDT, Sigma) by incubation at 37°C for 60 min and alkylated with 25 mM of S-methyl methanethiosulfonate (MMTS, Fluka) for 30 min at room temperature. One half of the sample was digested by 500 ng of lysyl endopeptidase (Lys-C, Fujifilm Wako Pure Chemical Corporation) and incubated for 5 hours at 25°C, before being diluted to 2 M urea with 100 mM ABC and being further digested with 500 ng trypsin (Trypsin Gold, Promega) at 37°C overnight. The second half of the sample was diluted to 1 M urea with 100 mM ABC and digested with 500 ng Chymotrypsin (Sequencing grade, Roche) for 5h at 25°C. The digests were acidified by addition of trifluoroacetic acid (TFA, Pierce) to 1%. A similar aliquot of each sample was analysed by LC-MS/MS.

#### LC-MS/MS

The Vanquish Neo UHPLC-System was coupled to an Orbitrap Exploris 480 mass spectrometer, equipped with a FAIMS pro interface and a Nanospray Flex ion source (all parts Thermo Scientific). Peptides were loaded onto a trap column (PepMap Acclaim C18, 5 mm × 300 μm ID, 5 μm particle size, 100 Å pore size, Thermo Scientific) at a flow rate of 25 μl/min using 0.1% TFA as mobile phase.

After 10 minutes, the trap column was switched in line with the analytical column (PepMap Acclaim C18, 500 mm × 75 μm ID, 2 μm, 100 Å, Thermo Fisher Scientific) operated at 30°C. Peptides were eluted using a flow rate of 230 nl/min, starting with the mobile phases 98% A (0.1% formic acid in water) and 2% B (80% acetonitrile, 0.1% formic acid) and linearly increasing to 35% B over the next 90 minutes.

The Orbitrap Exploris 480 mass spectrometer was operated in data-dependent mode, performing a full scan (m/z range 350-1200, resolution 60,000, normalized AGC target 300%) at 3 different compensation voltages (CV -45, -60, -75), followed each by MS/MS scans of the most abundant ions for a cycle time of 1.0 seconds per CV. MS/MS spectra were acquired using HCD collision energy of 30, isolation width of 1.2 m/z, orbitrap resolution of 30.000, normalized AGC target 200%, minimum intensity of 50.000 and maximum injection time of 100 ms. Precursor ions selected for fragmentation (include charge state 2-6) were excluded for 10 s. The monoisotopic precursor selection filter and exclude isotopes feature were enabled.

#### Data Processing protocol

For peptide identification, the RAW-files were loaded into Proteome Discoverer (version 2.5.0.400, Thermo Scientific). All MS/MS spectra were searched using MSAmanda v2.5^51^. The peptide mass tolerance and fragment mass tolerance was set to ±10 ppm, the maximum number of missed cleavages was set to 2, using tryptic or chymotryptic enzymatic (semi-)specificity without proline restriction. Peptide and protein identification was performed in two steps. The RAW-files were searched against the Trichoplusia Ni NCBI UniProt database (21163 sequences; 13598141 residues) and the baculovirus database (5310 sequences, 1749589 residues) supplemented with common contaminants and sequences of tagged protein of interest (351 sequences) using the following additional variable modifications: Carbamidomethylation and beta-methylthiolation on cysteine, oxidation on methionine, deamidation on asparagine and glutamine, phosphorylation on serine, threonine and tyrosine, ubiquitination on lysine, glutamine to pyro-glutamate conversion at peptide N-terminal glutamine and acetylation on protein N-terminus and carbamylation on lysine and the peptide n-terminus. The localization of the post-translational modification sites within the peptides was performed with the tool ptmRS, based on the tool phosphoRS^52^. Identifications were filtered to 1 % FDR on protein and PSM level, additionally an Amanda score cut-off of at least 150 was applied. Proteins were filtered to be identified by a minimum of 2 PSMs in at least 1 sample. Protein areas have been computed in IMP-apQuant^53^ by summing up unique and razor peptides. Resulting protein areas were normalized using iBAQ^54^ and sum normalization was applied for normalization between samples. Match-between-runs (MBR) was applied for peptides with high confident peak area that were identified by MS/MS spectra in at least one run.

## SUPPLEMENTARY FIGURES

**Figure S1.**
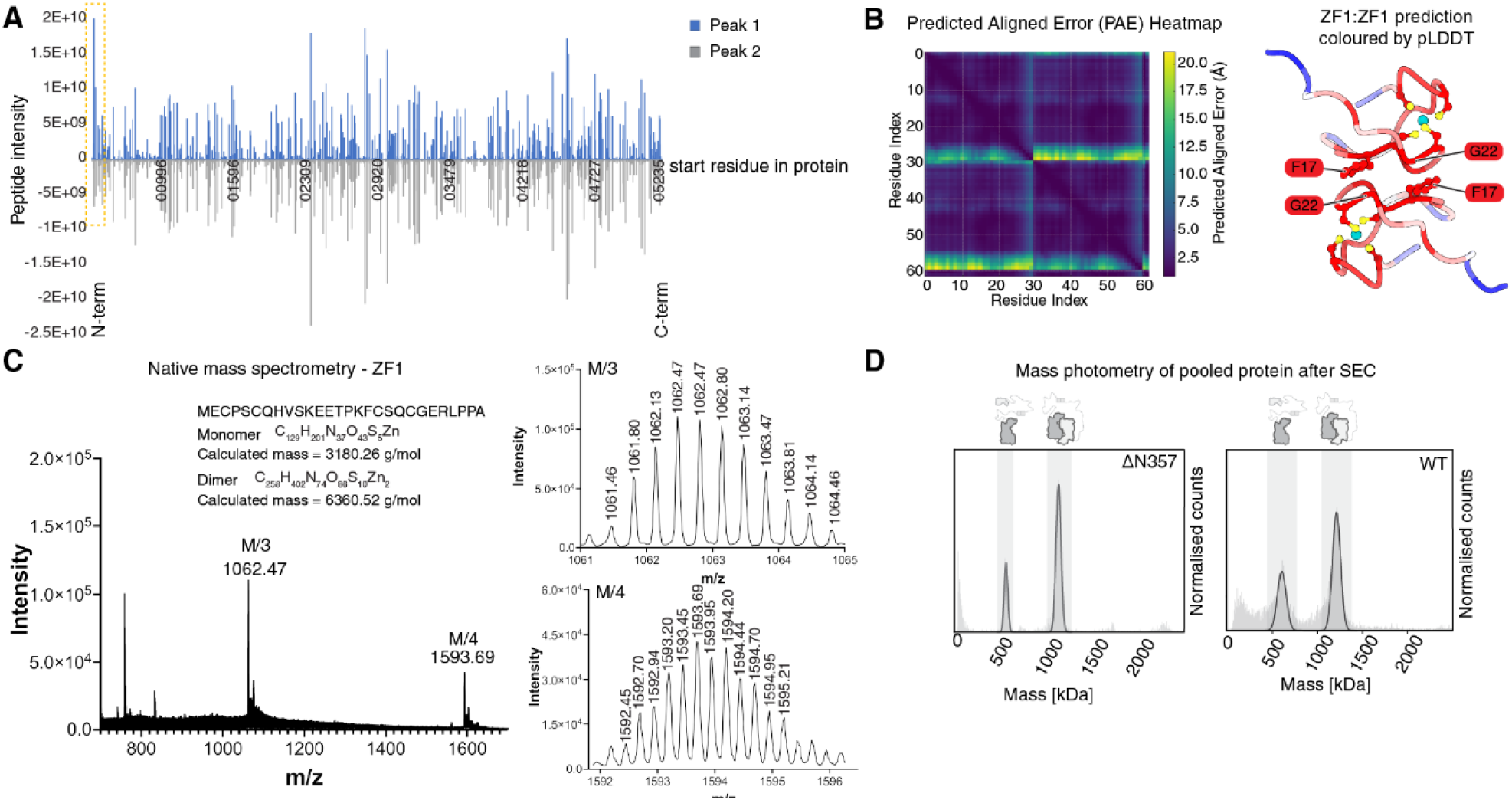
The N-terminal domain ZF1 of RNF213 is a high-affinity dimerization motif. **(A)** LC-MS/MS analysis of RNF213 WT peak 1 and 2 (related to **Figure 1A**). PSMs per detected peptide plotted for RNF213 from N-to C-terminus for Peak 1 and Peak 2. Yellow zone box indicates region where PSMs differ between Peak 1 and 2. **(B)** Predicted Aligned Error (PAE) plot for ZF1-ZF1 dimer (right; coloured by pLDDT) predicted by AF3 with an iPTM of 0.75, related to **Figure 1B. (C)** Analysis of ZF1 synthetic peptide by native mass spectrometry. Left; Full mass spectrum of ZF1 under native conditions with given sequence and calculated molecular weight for zinc-chelated monomer and dimer. Right, top; Zoomed in isotope pattern at peak 1062.47 corresponding to the monomer at charged state 3+ with zinc chelated. Right, bottom; Zoomed in isotope pattern at peak 1593.69 corresponding to the dimer at charged state 4+ with two zincs chelated. **(D)** Mass photometry analysis of recombinant RNF213 ΔN357 (where the N-terminal 357 residues were removed, including the N-terminal dimerising ZF1), compared to RNF213 WT (right).

**Figure S2.**
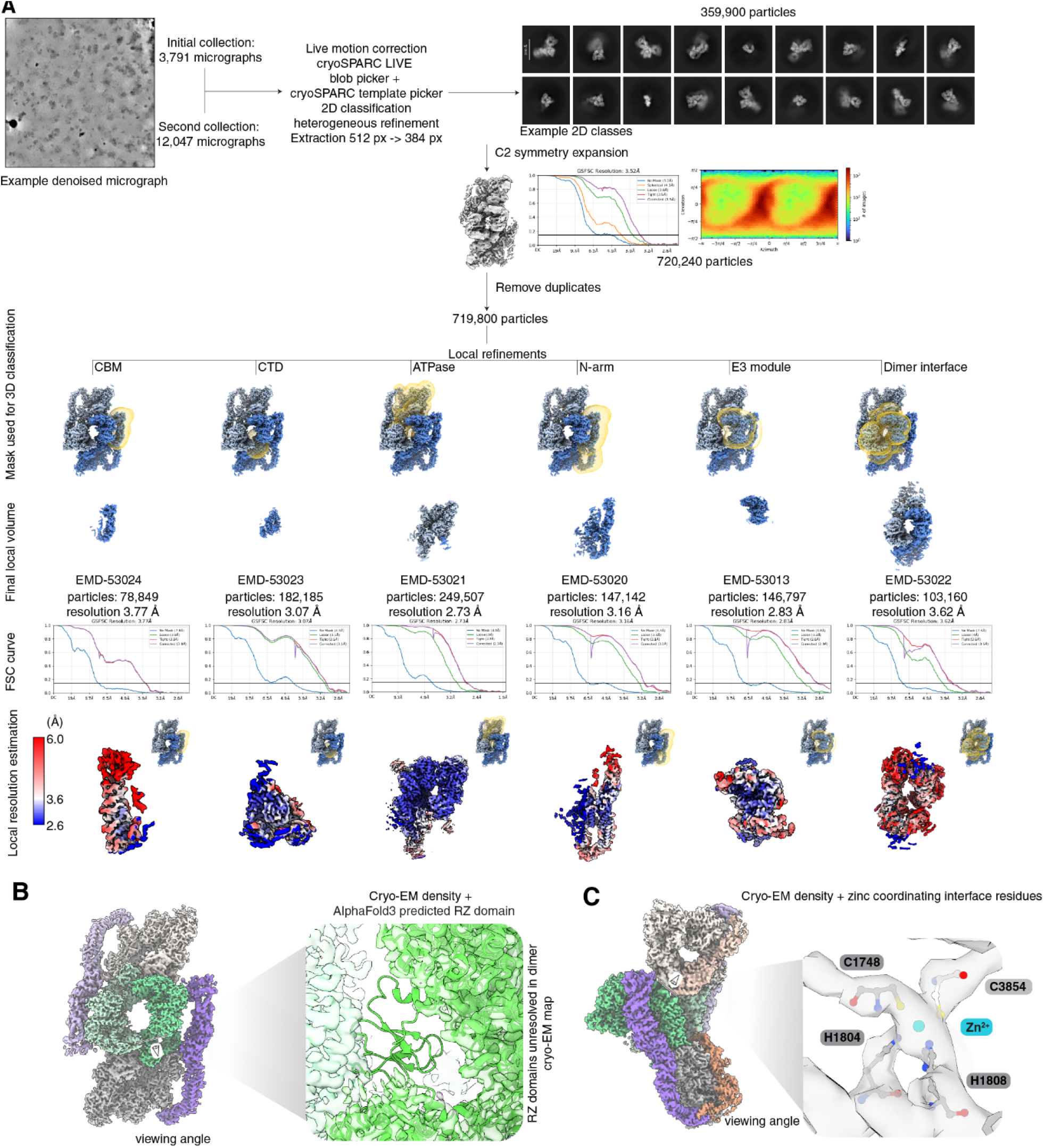
Cryo-EM workflow and structural features of the RNF213 closed dimer. **(A)** Cryo-EM processing pipeline for the hsRNF213 apo dimer. All additional cryo-EM datasets in this manuscript were processed in an identical manner until C2 symmetry expansion, at which point other datasets were not symmetry expanded before local refinements and 3D variability analysis. Additional information on the other datasets can be found in **Figure S5** and **Figure S8. (B)** Viewing angle (left) of zoomed (right) region displaying lack of visible density for RZ domain. Cryo-EM density and resulting model shown in shades of green and docked AlphaFold3 structure of the RZ domain aligned to draw attention to region of missing density. **(C)** Viewing angle (left) of zoomed (right) region of core dimer interface density and model. Residues C1748, H1804, H1808 and C3854 involved in zinc coordination displayed as ball-and-stick atoms.

**Figure S3.**
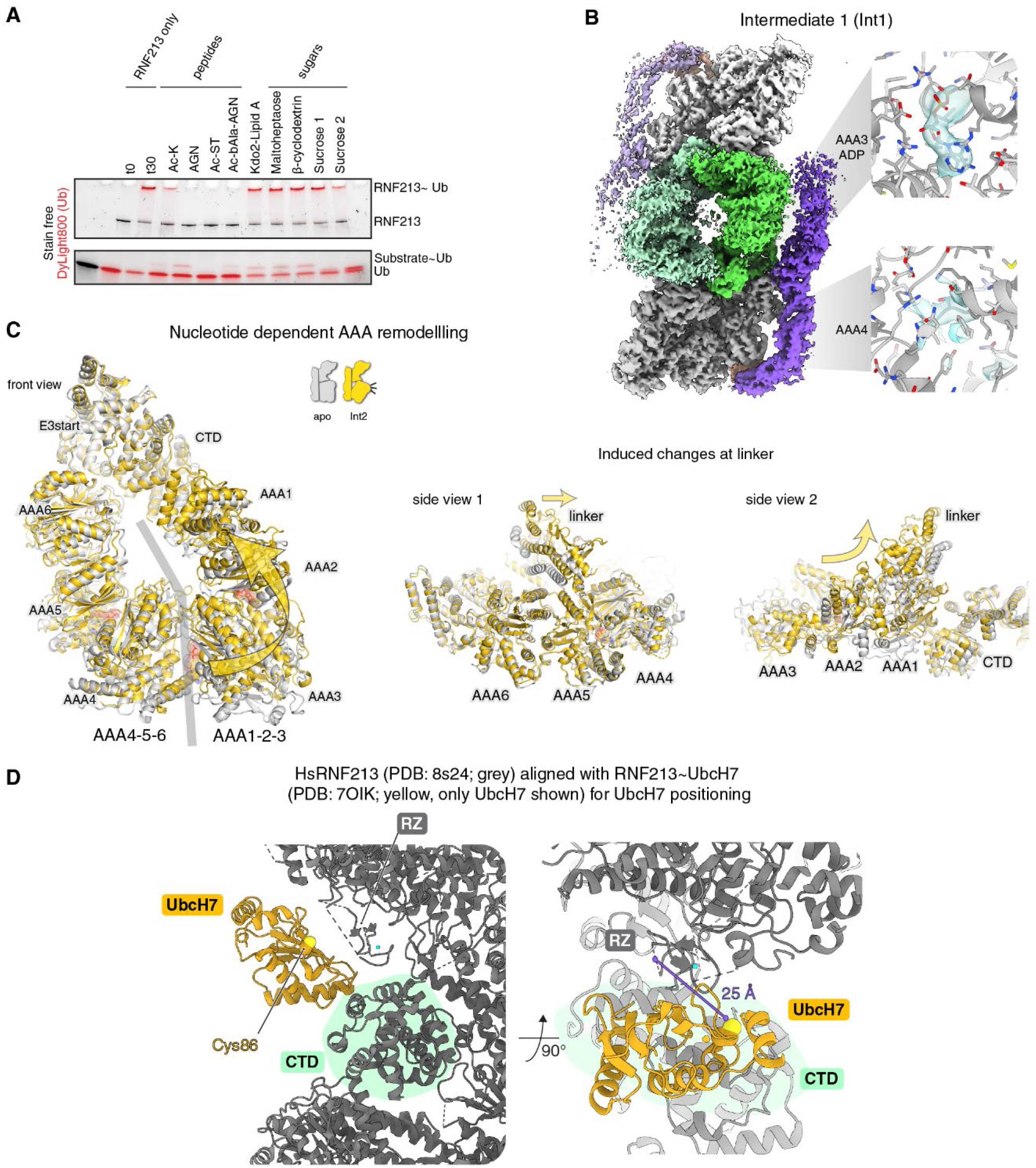
RNF213 nucleotide-dependent activity is stimulated through conformational remodelling. **(A)** Ubiquitination assay of RNF213 in the presence of ATP and either a peptide, lipid or sugar substrate as indicated above each lane. **(B)** Cryo-EM density of an intermediate of the open-closed transition (Int1), reflecting a state similar to the closed dimer structure. Here, a bound ADP was observed in the AAA3 active site, whereas no nucleotide was observed in AAA4 (right panel). (**C)** Superposition of the closed dimer (grey) with Int2 (gold), focusing on the AAA core and linker portions. Left: arrow indicates reorientation of the AAA1/2/3 half against the AAA4/5/6 half. Right: side views depicting rearrangement of the helical linker resulting from the 15° tilt of AAA1/2/3, leading to a disruption of the core interface near the Zn^2+^ site. **(D)** HsRNF213 monomer (PDB:8s24; grey) aligned with the RNF213 monomer bound to UbcH7 ABP (PDB: 7OIK) (monomeric form hidden, UbcH7 shown in yellow). Indicated distance is between the catalytic cysteines of UbcH7 and RZ of RNF213 as measured in ChimeraX.

**Figure S4.**
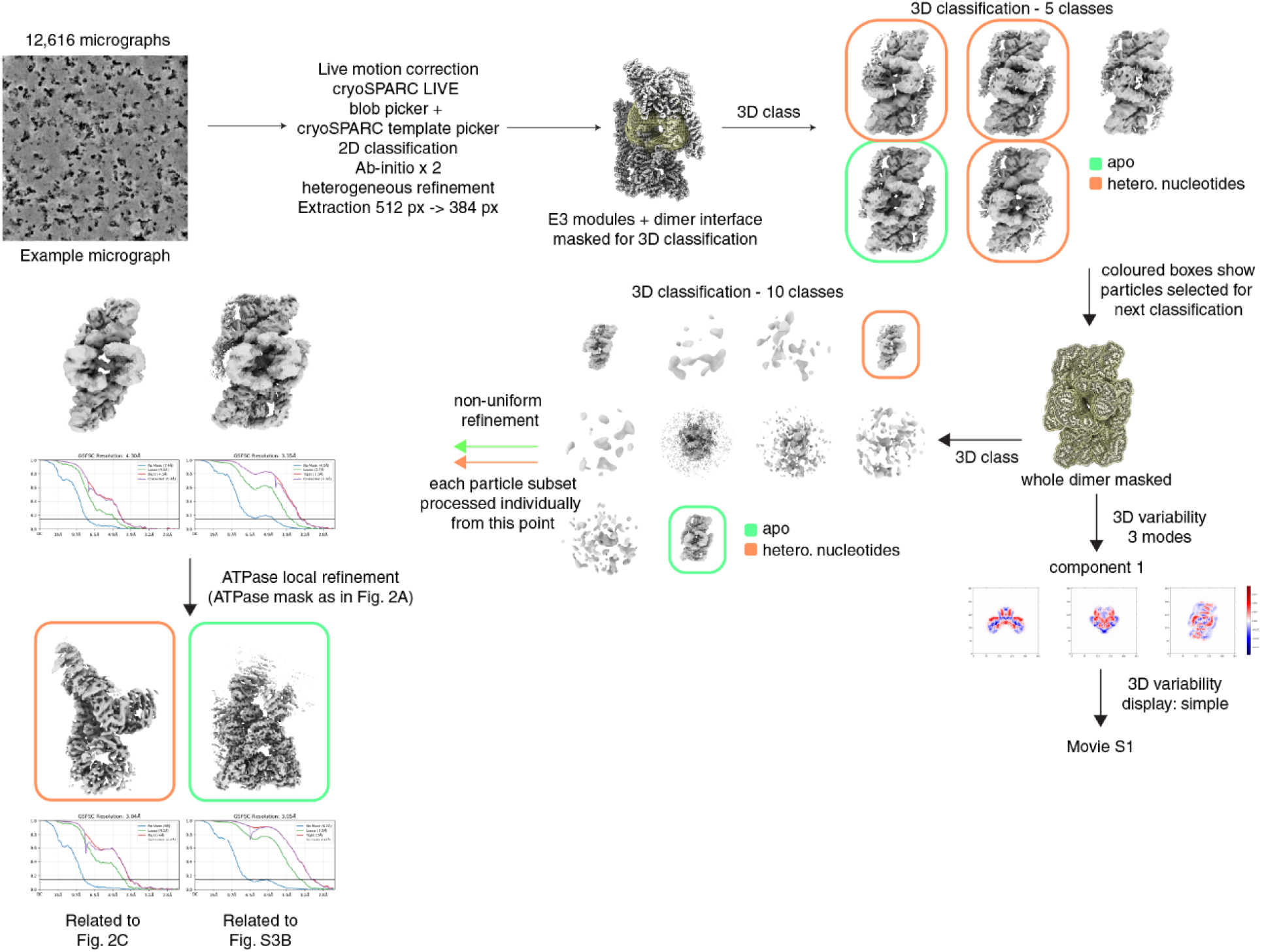
Cryo-EM processing workflow for ATPγS-treated RNF213. Cryo-EM processing pipeline for the hsRNF213 + ATPγS dataset. This figure highlights only the aspects of data processing that explicitly differ from that of the apo dataset found in **Figure S2A**.

**Figure S5.**
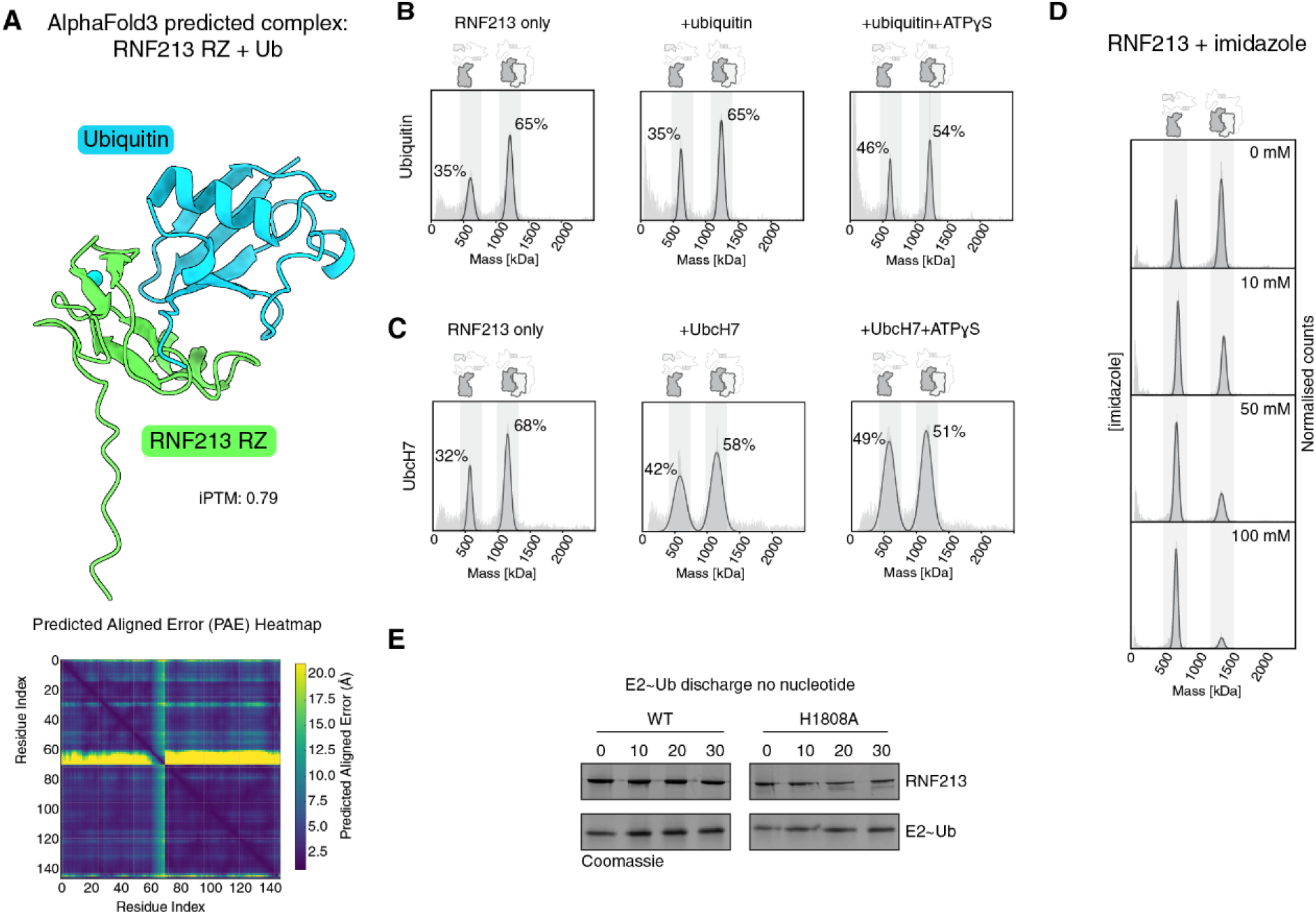
Ubiquitination factor binding and zinc chelation stabilise the open dimer conformation. **(A)** AlphaFold3 prediction of hsRNF213 RZ domain (residues 4486-4555; green) binding ubiquitin (blue) with iPTM = 0.79, alongside PAE plot of prediction **(B)** Single point mass photometry measurements of RNF213 only, RNF213+ubiquitin and RNF213+ubiquitin+ATPγS, where the y-axis shows normalised counts, and percentage contribution of each peak to the total counts of RNF213 species. Profiles were recorded after 30 min incubation. **(C)** Single point mass photometry measurements of RNF213 only, RNF213 + UbcH7 and RNF213 + UbcH7 + ATPγS, recorded and evaluated as in (B). **(D)** Mass photometry analysis of RNF213 incubated with an increasing titration series of imidazole including an RNF213 only control (top), followed by addition of imidazole from 10 to 100 mM. **(E)** E2∼Ub discharge assay using WT or H1808A RNF213 in the absence of activating nucleotide stimulus. Coomassie stained gels cropped to display the concentrations of RNF213 variants (top) and amount of E2∼Ub (bottom) remaining after 30 minutes.

**Figure S6.**
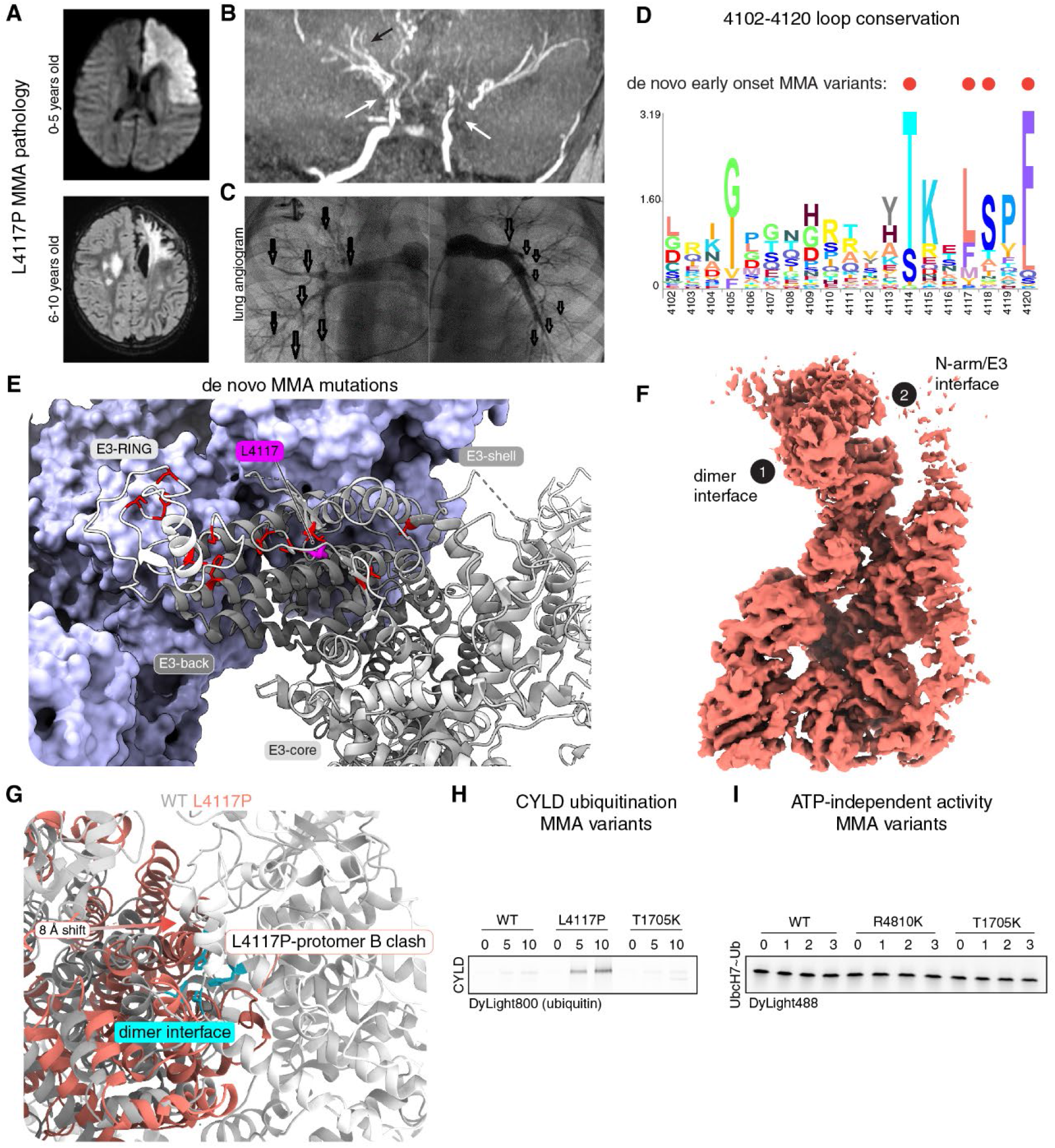
Moyamoya disease mutant pathology, structure and function. **(A)** Top: axial diffusion-weighted MRI from a patient carrying the RNF213 L4117P variant, obtained between 0-5 years of age, showing an acute infarct in the left anterior watershed territory. Bottom: axial FLAIR MRI from the same patient between 6-10 years of age showing progression of parenchymal ischemic damage with bilateral cortical and white matter lesions. **(B)** Coronal time-of-flight MR angiography from the same patient showing bilateral stenosis of the terminal internal carotid arteries (white arrows) and lenticulostriate collateral vessels (black arrow), characteristic of Moyamoya angiopathy. **(C)** Selective pulmonary angiography from the same patient showing well-developed proximal right and left pulmonary arteries (arrows) but severe hypoplasia of the distal pulmonary vascular tree bilaterally. **(D)** Conservation plot for residues 4102-4120 MMA mutant hotspot where larger letters illustrate greater conservation. Residues affected by de novo MMA variants are labelled with a red dot. **(E)** Zoomed-in view of de novo, early onset MMA variants (red), with the different E3 module regions (RING, back, core and shell) shown in different shades of grey and labelled. Residue L4117, identified as a recurrent site of MMA mutation in this study, is highlighted in magenta. **(F)** 3D reconstruction of L4117P RNF213 with insets 1 and 2 highlighted and shown in zoomed views in **Fig. 4E-F. (G**) RNF213 apo dimer (grey/white) overlaid with L4117P cryo-EM model (red) where L4117P and protomer A are aligned on their AAA modules. Dimer interface residues C1748, H1804, H1808 and C3854 are highlighted on the apo dimer in blue. Arrows indicate movement of L4117P relative to protomer A, and resulting clash that would occur with protomer B. **(H)** CYLD ubiquitination assay showing that L4117P has increased substrate ubiquitination activity compared to WT, but T1705K performs comparably to WT. Numbers above gel indicate timepoints taken in minutes. **(I)** E2∼Ub discharge is not permitted in MMA variants R4810K and T1705K in the absence of activating nucleotide, as compared to WT RNF213. Numbers indicate timepoints taken in hours.

**Figure S7.**
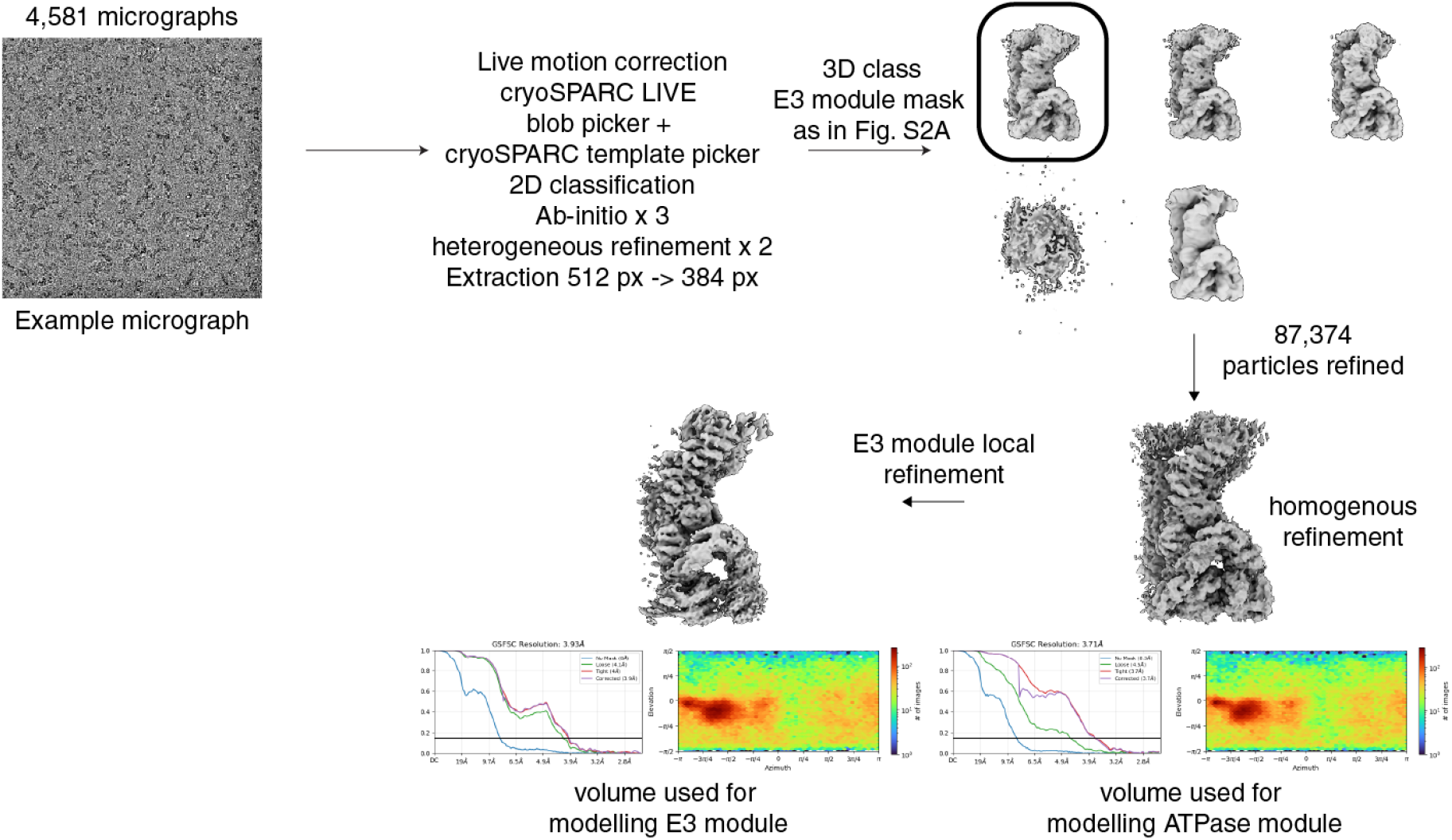
Cryo-EM processing workflow for the L4117P RNF213 mutant. Cryo-EM processing pipeline for the L4117P RNF213 dataset. This figure highlights the aspects of data processing that explicitly differ from that in **Figure S2A**. Note that when modelling RNF213 into the final volumes presented here, the RING domain was excluded owing to the heterogeneity of the density which made it unclear where the domain would be positioned.

**Figure S8.**
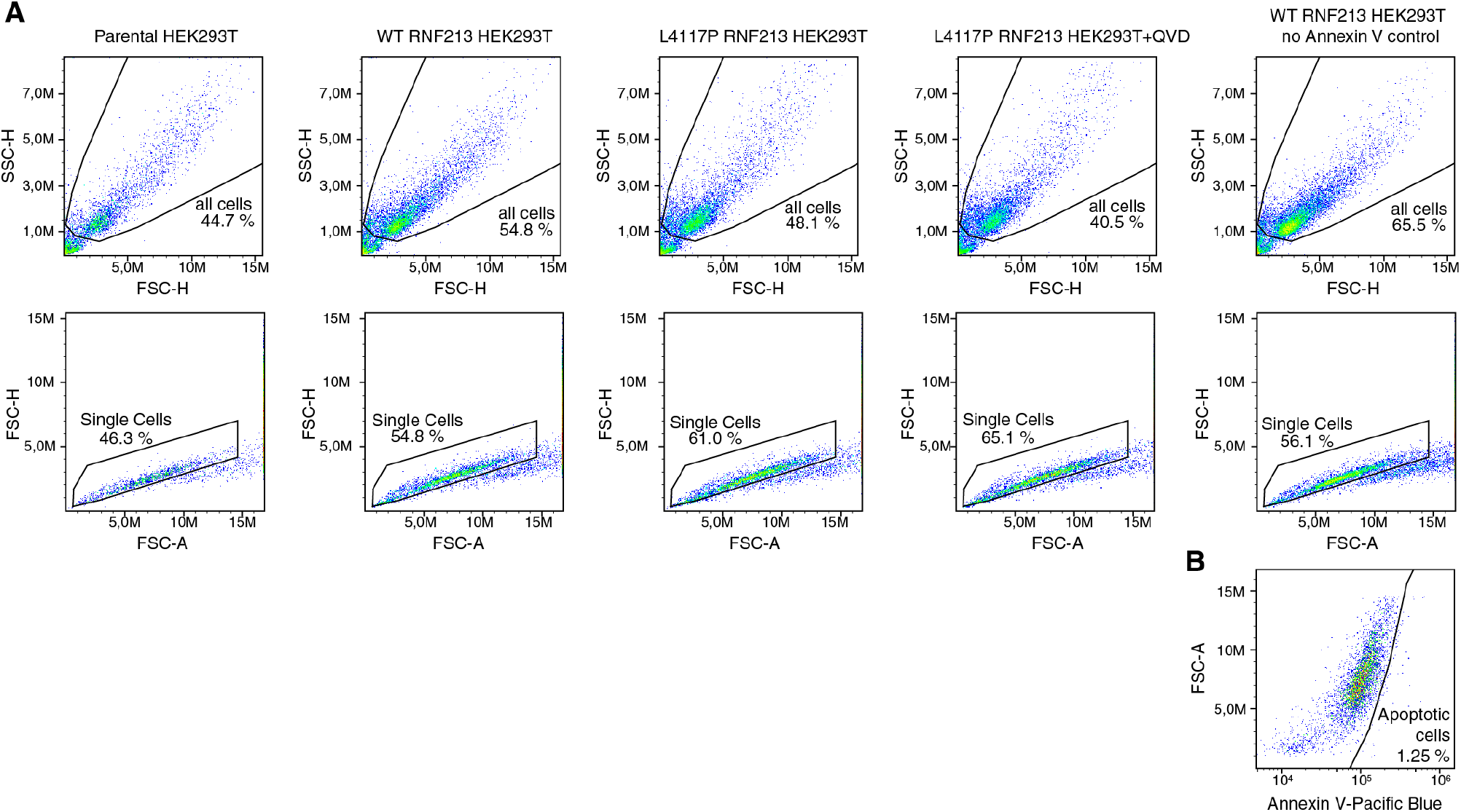
FACS analysis of parental and inducible RNF213 HEK293T cell lines. **(A)** Gating parameters for all cells and live cells for FACS analysis, prior to apoptosis gating shown in **Figure 5H. (B)** Gating for apoptotic cells selected by first measurement of WT RNF213 HEK293T cells analysed by FACS but not stained with Annexin V.

## SUPPLEMENTARY TABLES

**Table S1.**
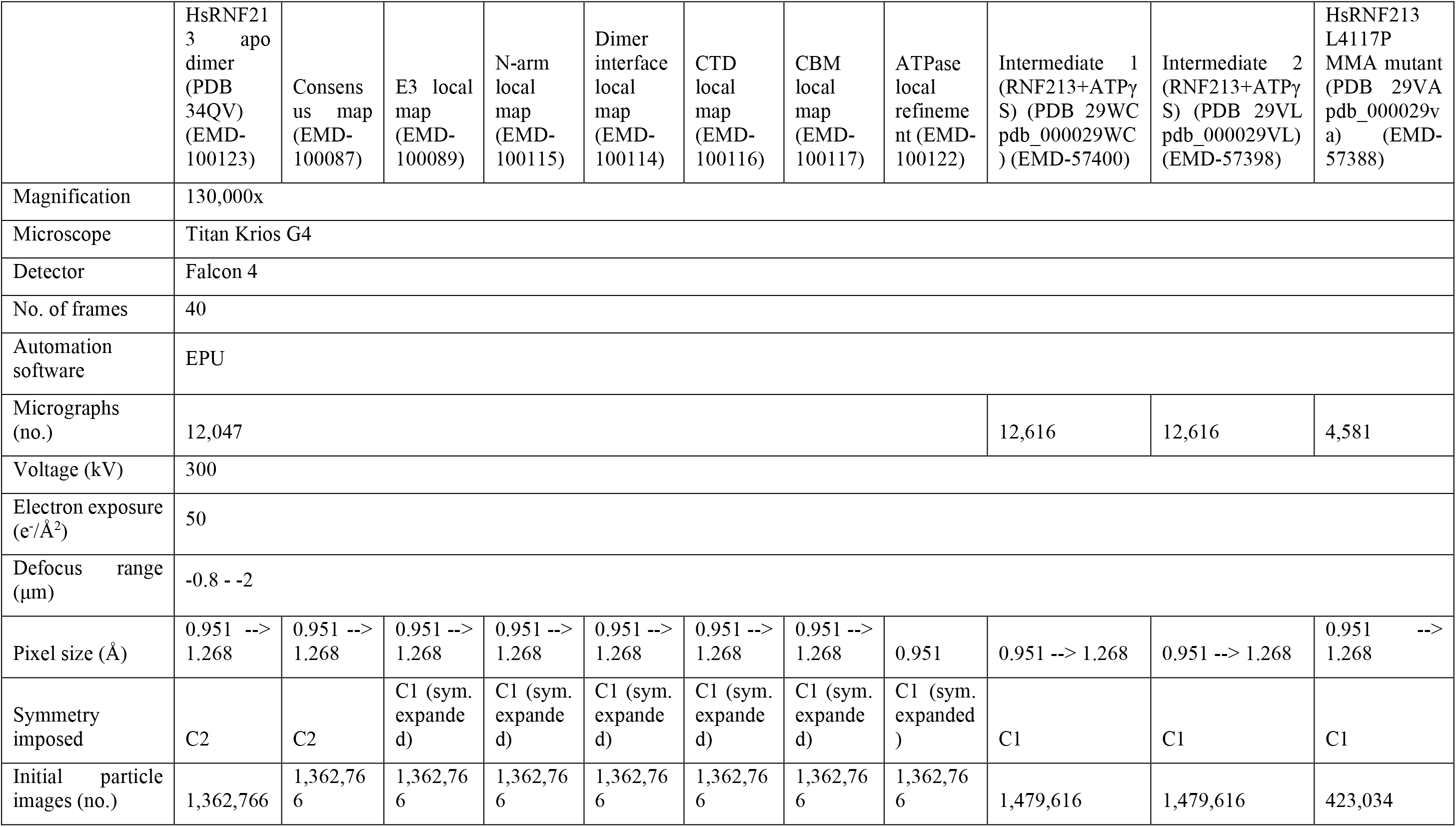
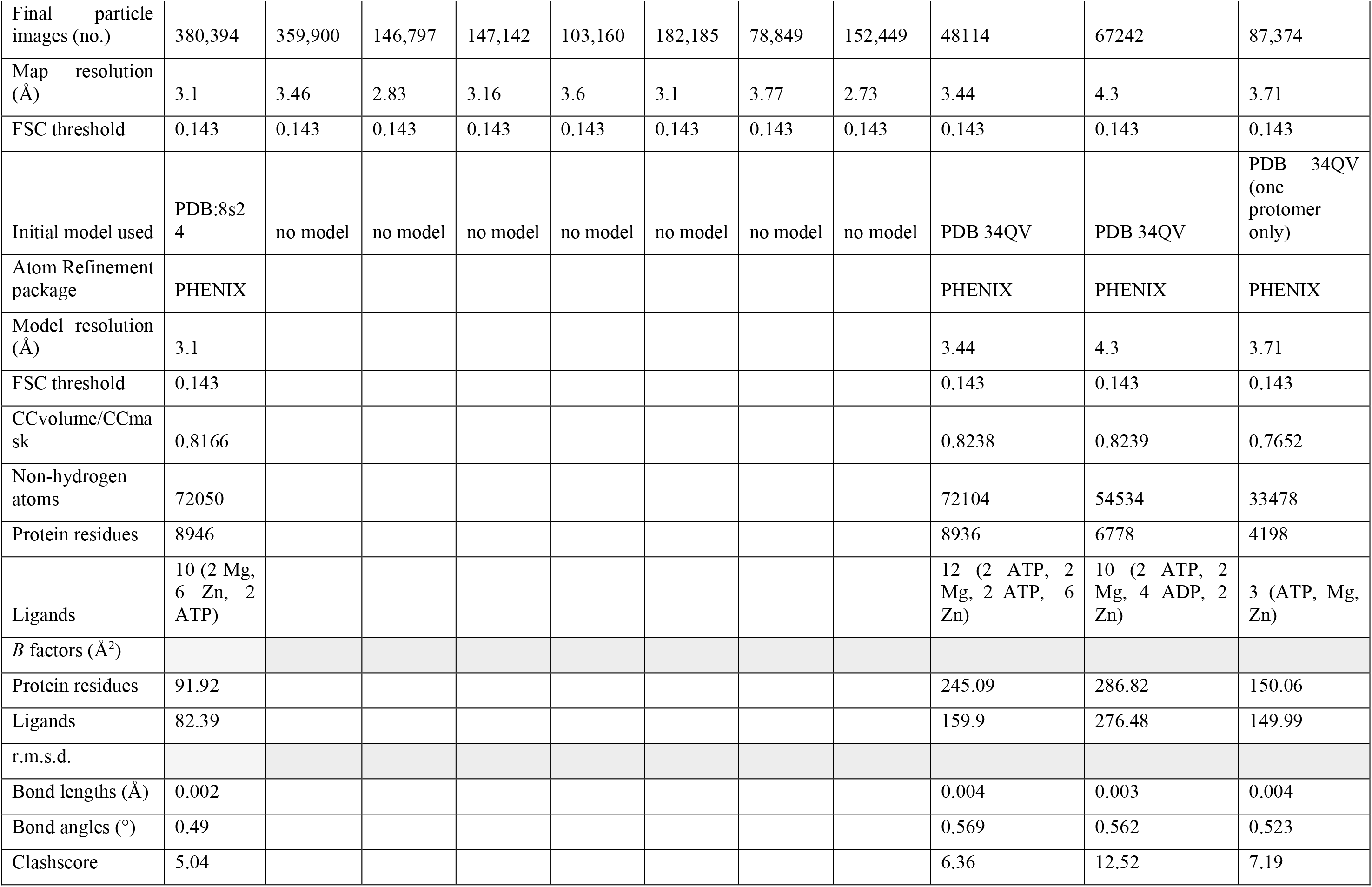
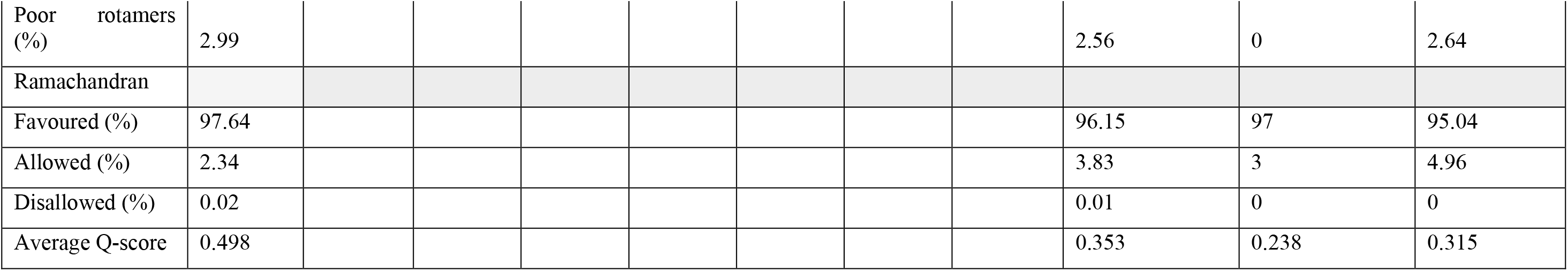
Cryo-EM data collection and refinement statistics.

**Table S2.** CADD score (h38-v1.6) of selected RNF213 MMA variants.

| Variant | CADD |
| --- | --- |
| **L4117P\*\*** | **28.0** |
| L4117R | 27.6 |
| F4120L | 27.5 |
| S4118F | 27.1 |
| R4062Q | 27.0 |
| V4941E | 25.7 |
| T4114R | 25.7 |
| C4032R | 25.5 |
| C4012R | **25.4** |
| C4035R | 25.1 |
| C3997G | **25.1** |
| C3997Y | 25.0 |
| V4146A | 24.9 |
| H4058P | 24.8 |
| C4017S | **24.7** |
| K4185E | 24.5 |
| D4237E | 21.7 |
| **T1705K\*\*** | **23.5** |
| H4014N | **23.3** |
| P4007R | 23.3 |
| K4185T | 23.2 |
| S1474F | 22.9 |
| V4765M | 22.5 |
| P1721L | 20.9 |
| D4013N | 20.3 |
| A1844T | 19.1 |
| Q4367L | 16.7 |
| R4019C | 15.5 |
| A5021V | 15.4 |
| P4608S | 15.0 |
| **R4810K\*\*** | **7.4** |

residues in **bold** – de novo, early-onset MMA variants; remaining MMA variants extracted from Ahel et al (2020) with CADD scores > 15 plus the founder variant R4810K.
**\*\*** variants used for comparative functional studies, having high (L4117P), medium (T1705K) and low (R4810K) CADD scores

**Table S3.** De novo early-onset, severe RNF213 mutations identified here and in recent work.

| **Variant** | **Phenotype** | **Reference** |
| --- | --- | --- |
| **L4117P** | stroke events starting at 0-5 years old, liver transaminases increase, pulmonary arteries stenosis | present work |
| **L4117P** | liver disease, bilateral stenosis of intracranial arteries typical of MMA | present work |
| **L4117R** | Recurrent strokes at 0-5 years old, liver cytolysis and kidney disease | present work |
| **F4120L** | stroke events starting at 0-5 years old, pulmonary arteries hypertension, stenosis of left renal artery | present work |
| **K4136Q** | First stroke at 0-5 years old, severe hypertension, stenosis of the right renal artery | present work |
| **E4942-V4948 (duplication)** | stroke events starting at 0-5 years old, liver cholestasis detected, hypogammaglobulinemia | present work |
| **P3996-C3997 (replaced with GLG)** | 1st stroke at 0-5 years old, liver cytolysis, bilateral renal arteries stenosis, occlusion of the iliac artery | 55 |
| **C4017S** | MMA, no specific clinical data | 56 |
| **H4058P** | Cerebral infarction at 0-5 years old | 57 |
| **T4114R** | severe childhood moyamoya/diffuse vasculopathy | 55 |
| **S4118F** | First stroke at 0-5 years old, liver cytolysis | 58 |
| **S4118C** | MMA, liver cytolysis, stenosis of extracerebral arteries | 59 |
| **F4120L** | onset 0-5 years old | 55 |
| **F4120L** | MMA, liver cytolysis, stenosis of extracerebral arteries | 59 |
| **L4139W** | First stroke at 0-5 years old, liver cytolysis, erythema and kidney disease | 29 |
| **T4155P** | Cerebral infarction and haemorrhage at 0-5 years old | 57 |
| **C4856R** | First stroke at 0-5 years old, erythema, kidney disease | 29 |

## Notes

### Competing Interest Statement

The authors have declared no competing interest.

### Author Declarations

Since mutation screening was performed in a diagnostic context it does not require in France an IRB approval (Molecular testing is not performed in a research project but for diagnostic). The required consent forms for diagnostics and for use of their data for research purposes were signed by either the patient or both parents in case of an affected child. All patients for which data has been included in this study have been anonymised.

